# Characterizing cell type specific transcriptional differences between the living and postmortem human brain

**DOI:** 10.1101/2024.05.01.24306590

**Authors:** Eric Vornholt, Lora E. Liharska, Esther Cheng, Alice Hashemi, You Jeong Park, Kimia Ziafat, Lillian Wilkins, Hannah Silk, Lisa M. Linares, Ryan C. Thompson, Brendan Sullivan, Emily Moya, Girish N. Nadkarni, Robert Sebra, Eric E. Schadt, Brian H. Kopell, Alexander W. Charney, Noam D. Beckmann

## Abstract

Single-nucleus RNA sequencing (snRNA-seq) is often used to define gene expression patterns characteristic of brain cell types as well as to identify cell type specific gene expression signatures of neurological and mental illnesses in postmortem human brains. As methods to obtain brain tissue from living individuals emerge, it is essential to characterize gene expression differences associated with tissue originating from either living or postmortem subjects using snRNA-seq, and to assess whether and how such differences may impact snRNA-seq studies of brain tissue. To address this, human prefrontal cortex single nuclei gene expression was generated and compared between 31 samples from living individuals and 21 postmortem samples. The same cell types were consistently identified in living and postmortem nuclei, though for each cell type, a large proportion of genes were differentially expressed between samples from postmortem and living individuals. Notably, estimation of cell type proportions by cell type deconvolution of pseudo-bulk data was found to be more accurate in samples from living individuals. To allow for future integration of living and postmortem brain gene expression, a model was developed that quantifies from gene expression data the probability a human brain tissue sample was obtained postmortem. These probabilities are established as a means to statistically account for the gene expression differences between samples from living and postmortem individuals. Together, the results presented here provide a deep characterization of both differences between snRNA-seq derived from samples from living and postmortem individuals, as well as qualify and account for their effect on common analyses performed on this type of data.

## INTRODUCTION

In recent years, single-nucleus RNA sequencing (snRNA-seq), which quantifies transcript abundance across thousands of genes within nuclei from individual cells in a single tissue sample^1^, has become a preferred approach to studying human brain biology. Applications of snRNA-seq for human brain biology research include: (1) defining gene expression patterns characteristic of brain cell types and states^2,3^, (2) identifying differences in gene expression for a given cell type between two groups of brain samples (e.g., cases and controls)^4–7^, and (3) estimating the proportions of different brain cell types present in bulk RNA-seq data^8,9^, which represents transcript abundance from RNA pooled across hundreds to millions of heterogeneous cells in a single homogenized sample^10^. These applications of snRNA-seq have resulted in large databases (i.e., atlases) characterizing the diversity of cell types in multiple brain regions^11,12^ as well as insights into the cell types and gene expression patterns underlying brain disorders^13–16,17–19^.

Since it is difficult to obtain brain tissue for research from living individuals, most applications of snRNA-seq to study human molecular neurobiology have used brain tissue obtained from postmortem donors. Recently, several research initiatives have highlighted the value of studying brain tissue from living individuals^12,20–24^, including the Living Brain Project (LBP). In a companion LBP report by Liharska *et al.*^23^, molecular differences between living and postmortem brain tissue are characterized using bulk RNA-seq to show greater than 80% of all genes analyzed were differentially expressed between living and postmortem samples^23^. However, neither Liharska *et al.* nor the other recent studies utilizing brain tissues from living individuals have adequately explored transcriptional differences between samples obtained from living and postmortem individuals at a single-nuclei resolution.

Here, expression differences between living and postmortem brain samples are identified through analyses of snRNA-seq data generated from living and postmortem PFC samples (***Figure 1a***). The primary findings of this report are: (1) the same cell types are identified in living and postmortem nuclei, (2) there is a differential expression signal between living and postmortem nuclei across cell types, (3) estimation of cell type proportions by cell-type deconvolution is more accurate in living samples than in postmortem samples, and (4) differences between living and postmortem brains can be statistically accounted for in snRNA-seq and bulk RNA-seq data. Together, these results help further characterize the molecular differences between living and postmortem samples, as well as qualify transcriptional differences on common snRNA-seq and bulk RNA-seq based approaches to study human neurobiology.

**Figure 1:**
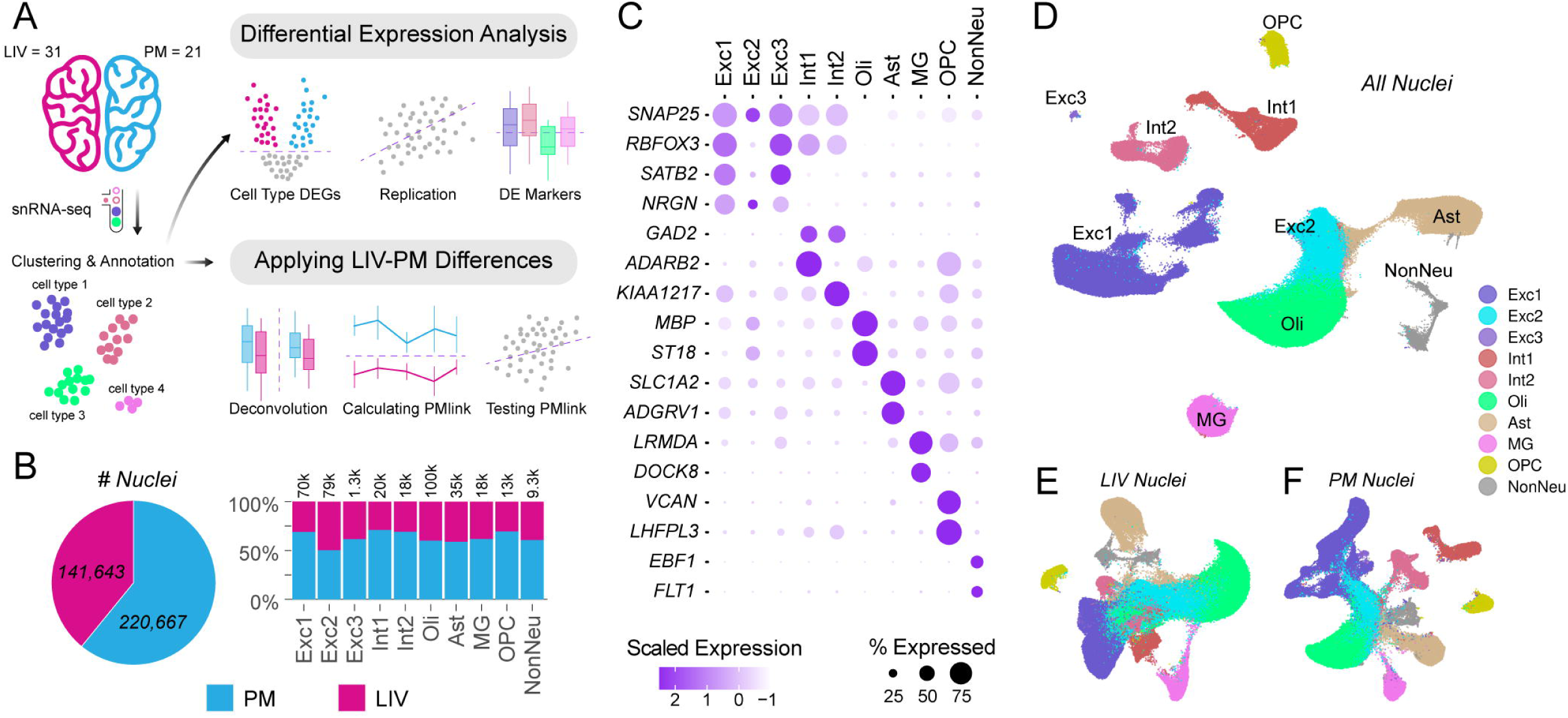
Study overview and snRNA-seq clustering. ***A)*** Overview of study design. Nuclei from living (LIV=31) and postmortem (PM=21) samples were sequenced using snRNA-seq. After QC, nuclei were clustered and annotated into cell types. For each cell type, differential expression (DE) analyses were performed to identify genes with increased abundance in LIV sample and genes with increased abundance in PM samples. DE signatures were replicated and the effect of the LIV-PM status on cell type marker genes was defined. Annotated nuclei were used to establish differences in deconvolution accuracy between LIV and PM mixtures using LIV or PM references. A continuous metric (PMlink) capable of accounting for differences in gene expression between LIV and PM samples was developed and characterized. ***B)*** Post-QC LIV (pink) and PM (blue) nuclei counts/proportions for the entire cohort for all nuclei (left, pie chart) and for each cell type (right, barplot). The x and y axes of the barplot represent respectively cell type and percent of nuclei present in the cohort for that cell type. The total number of nuclei for each cell type is defined on the top of each bar. ***C)*** Dotplot of the expression of selected canonical markers used for cell annotation. The x and y axes are cell type and the union of the top 2 gene markers for each cell types. Dot size represents the percentage of cells in a specific cluster expressing the marker and color intensity represents the scaled expression of each marker. ***D)*** Uniform manifold approximation and projection (UMAP) coordinates for all-nuclei colored and annotated based on cell type identity as defined in the legend. ***E)*** UMAP coordinates for clustering performed on only LIV nuclei colored based on cell type classifications from the all-nuclei annotations as defined in the legend. ***F)*** UMAP coordinates for clustering performed on only PM nuclei colored based on cell type classifications from the all-nuclei annotations as defined in the legend.

### Living Brain Project cohort

The biopsy procedures for obtaining living PFC tissue for the LBP are described in Liharska *et al*^23^. In brief, PFC samples from living participants are obtained during deep brain stimulation (DBS) surgery, an elective neurosurgical treatment for neurological and mental illnesses^23^. For the current report, a total of 33 PFC biopsies (“LIV samples”) from 23 living participants were used, including bilateral biopsies from 12 participants and unilateral biopsies from 9 participants (7 from the left hemisphere and 2 from the right hemisphere). For comparison, a cohort of postmortem PFC samples (“PM samples”, N = 23) was assembled from two brain banks (the Harvard Brain Tissue Resource Center and New York Brain Bank at Columbia University). To the extent possible, PM samples were matched to LIV samples for age and sex, and the majority of samples were obtained from individuals with Parkinson’s disease (PD), the most common indication for DBS (***Supplemental Table 1***).

Following library preparation, snRNA-seq was performed at the Icahn School of Medicine at Mount Sinai in New York City. The quality control procedures applied to the resulting snRNA-seq data described fully in the Methods take into consideration a variety of metrics characterizing the quality of both samples (e.g., number of nuclei) and nuclei (e.g., sequencing depth, percentage of transcripts originating from mitochondria, and detection of doublets). After applying filters based on these metrics, a total of 362,390 nuclei were retained for analysis from 52 samples, including 141,643 nuclei from 31 LIV samples (“LIV nuclei”) and 220,667 nuclei from 21 PM samples (“PM nuclei”) (***Figure 1b***). Hereafter, these 362,390 nuclei will be referred to as “the full LBP snRNA-seq nuclei set” and the 52 samples will be referred to as “the full LBP snRNA-seq cohort.”

### Cell types are conserved between living and postmortem human brain tissues

The same primary neural cell types were hypothesized to be present in human PFC samples regardless of whether a PFC sample is from a living participant or a postmortem donor (i.e., the “LIV-PM status” of a sample). To test this hypothesis, the following procedure was performed: (1) all nuclei were assigned to a cell type; (2) for each cell type, cell type markers (i.e., a group of genes differentiating that cell type from the other cell types) were defined for either LIV nuclei (“LIV markers”) or PM nuclei (“PM markers”) (***Supplemental Table 2***); (3) for each nucleus, two scores were calculated to capture the expression of the markers of its assigned cell type identity for LIV markers (“LIV marker scores”) and for PM markers (“PM marker scores”); (4) LIV marker scores were compared with PM marker scores (Spearman’s correlation). A total of 10 cell types were identified: three types of excitatory neurons (Exc1, Exc2, and Exc3), two types of inhibitory neurons (Int1 and Int2), oligodendrocytes (Oli), astrocytes (Ast), microglia (MG), oligodendrocyte progenitor cells (OPC), and non-neural cells (NonNeu; omitted from most downstream analyses) (***Figure 1b & c***). Both LIV nuclei and PM nuclei were well represented within each cell type (***Figure 1e & f*; *Supplemental Figure 1a & b***). The high correlation observed between LIV marker scores and PM marker scores (Spearman’s ρ = 0.93, p-value ≈ 0) confirms that the same main cell types which are identified by the same marker genes are present in human PFC samples regardless of the LIV-PM status of a given sample.

### Ubiquitous cell type specific differential expression between LIV and PM samples

With Liharska *et al.*^23^ identifying a large number of genes differentially expressed between LIV and PM samples in bulk RNA-seq data, analyses were performed to test the hypothesis that a similar pattern of differential expression (DE) would emerge when comparing LIV samples and PM samples for each of the 9 neural cell types identified above. The “pseudo-bulk” expression level (i.e., the sum of all counts for a gene across all cells from a sample) of every gene expressed in a given cell type was tested for association with LIV-PM status. Sex, PD status (i.e., whether a sample is from an individual with PD or not), and several additional technical metrics were used as covariates to account for unwanted variation in all statistical models described here. The log fold-change (logFC) calculated for each regression model captures both the magnitude and direction of the association between LIV-PM status and pseudo-bulk gene expression levels. The 9 resulting LIV-PM DE signatures (i.e., summary statistics for all of the genes tested in a specific cell type) are provided in ***Supplemental Table 3***. The percentage of significant differentially expressed genes (DEGs, FDR ≤ 0.05) was 26% for Exc1, 23% for Exc2, 37% for Exc3, 7% for Int1, 22% for Int2, 42% for Oli, 25% for MG, 19% for Ast, and 15% for OPC (***Figure 2a***). For each cell type, the percentage of DEGs that were “LIV DEGs” (i.e., DEGs more highly expressed in LIV samples) was generally similar to the percentage of “PM DEGs” (i.e., DEGs more highly expressed in PM samples) (proportion of DEGs that LIV DEGs: 50% for Exc1, 43% for Exc2, 48% for Exc3, 50% for Int1, 50% for Int2, 47% for Oli, 46% for MG, 50% for Ast, and 46% for OPC; ***Figure 2a*, *Supplemental Table 3***). Using the π_1_statistic^25^, which provides a lower bound of the percentage of genes tested that deviate from the null hypothesis of no association between gene expression and LIV-PM status, it was estimated that depending on the cell type, between 40% and 70% of genes are likely to be differentially expressed, and that the expected number of DEGs is not directly related to the number of nuclei in the cluster assessed (***Supplemental Figure 1c)***. Of the 27,316 genes expressed in at least one of the 9 neural cell types, 16,692 genes (61%) were found to be a DEG in at least one cell type, of which 57% were identified as DEGs in greater than one cell type (***Supplemental Figure 2***). Together, these results highlight cell type specific gene expression patterns associated with LIV-PM status in this LBP cohort.

**Figure 2:**
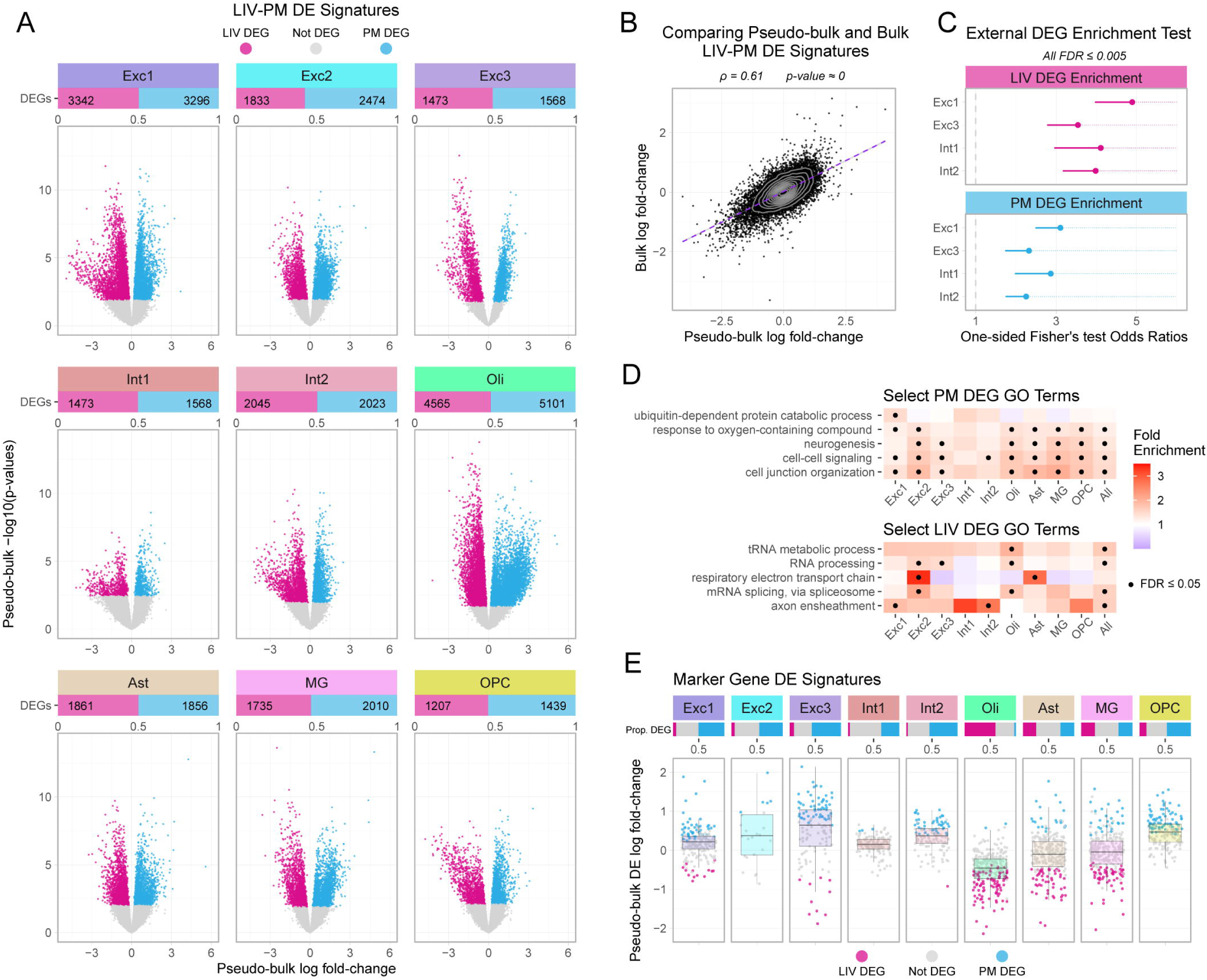
Differential expression between living and postmortem pseudo-bulk. ***A)*** Volcano plots presenting cell specific pseudo-bulk DE between LIV and PM samples. The x-axis represents the log fold-change (logFC) of the differential expression, the y-axis the −log10(p-value) of the association, and the color of each point represents the direction of association of each gene with the phenotype assessed as defined in the legend. The cell type assessed is defined above each plot. The proportion and number of significant DEGs (FDR ≤ 0.05) that are either LIV DEGs (higher transcript abundance in LIV; pink) or PM DEGs (higher transcript abundance in PM; blue) is presented above each volcano plot. ***B)*** Spearman’s correlation coefficient (ρ) between all-nuclei pseudo-bulk LIV-PM DE logFC and replication bulk LIV-PM DE logFC. The dashed purple line is best fitted line, and the grey contours represent the density of points. The ρ and associated p-value are defined above the plot. ***C)*** Plot of the replication of LIV-PM DEGs in external validation data. The x-axis is the odds ratios for the one sided Fisher’s exact tests of enrichment of LBP DEGs from cell types expressing RBFOX3 that overlap with DEGs from the Hodge et al. study, the y-axis is the cell types expressing RBFOX3, and the lines the 95% confidence interval. Each colored faced represents the enrichments of LIV DEGs (pink) and PM DEGs (blue). All odds ratios are significant with a FDR ≤ 0.005. ***D)*** Heatmap of biological processes gene ontology (GO) fold enrichment (low = blue, high = red) for LIV DEGs and PM DEGs from cell type and all-nuclei DE analyses with significant GO enrichment adjusted for multiple testing. The x-axis is the cell type, the y-axis selected representative significant GO term associated to the LIV-PM DEGs, the color is the fold enrichment as defined in the legend, the presence of a dot in each block represent significance (FDR ≤ 0.05), and the facets represent GO terms enriched for PM DEGs (top) and LIV DEGs (bottom). ***E)*** Boxplots of cell type markers DE for LIV-PM status. The facets represents different cell types, the y-axis is the logFC for the marker’s DE for LIV-PM status, and the color of point represents significance as defined in the legend. The proportion of marker genes from each cell type differentially expressed is defined above each box-plot with color representing direction of association as defined in the legend.

### PD status does not meaningfully impact LIV-PM DE signatures

Given the neurosurgical procedure where LIV samples were biopsied is often performed for the treatment of PD, the percentage of living participants diagnosed with PD (84%) was higher than the percentage of postmortem donors diagnosed with PD (48%). To ensure that this imbalance was not impacting the identification of DEGs associated with LIV-PM status, the samples in this cohort were subset to a PD only cohort (26 LIV samples and 10 PM samples) in which the cell type specific LIV-PM DE analyses were repeated. The concordance between the resulting PD-only LIV-PM DE signatures and the LIV-PM DE signatures described above (Spearman’s ρ range = 0.88-0.97, FDR ≈ 0); ***Supplemental Table 4***) suggests that these signatures are not explained by the imbalance of individuals with PD. This finding is consistent with observations made in Liharska *et al*^23^.

### LIV-PM differential expression replicates in independent snRNA-seq dataset

To assess the reproducibility of the LIV-PM DE signatures, the overlap was assessed between the DEGs identified here and DEGs from two independent studies that also compared gene expression between LIV and PM human brain samples: (1) the companion LBP study by Liharska *et al.*^23^ and (2) the study by Hodge *et al.*^12^. In Liharska *et al.*^23^, widespread differences in gene expression were identified in bulk RNA-seq data from PFC samples obtained for the LBP^23^. The bulk RNA-seq LIV-PM DE signature was compared to a LIV-PM DE signature generated on pseudo-bulk generated from all nuclei. The concordance between these two DE signatures (Spearman’s ρ = 0.61, p-value ≈ 0, ***Figure 2b***) demonstrates the reproducibility of the LIV-PM signatures identified here in bulk RNA-seq data.

Hodge *et al.*^12^ identified approximately 700 DEGs when comparing neuronal gene expression in the middle temporal gyrus (MTG) from four living individuals to neuronal gene expression in the MTG of four postmortem donors using snRNA-seq on nuclei positive for the neuronal marker NeuN. Hodge *et al.* DEGs were compared to the LIV-PM DEGs in the primary snRNA-seq LIV-PM DE signatures across all cell types that express RBFOX3, the gene encoding the NeuN protein (i.e., Exc1, Exc3, Int1, and Int2; ***Supplemental Figure 3***), DEGs with higher expression in PM samples relative to LIV samples in the Hodge *et al.* study significantly enriched for PM DEGs (ORs 2.25 – 3.11; all FDR < 7.03e^-6^) and the DEGs with higher expression in LIV samples relative to PM samples in the Hodge *et al.* study significantly overlapped with LIV DEGs (ORs 3.54 – 4.89; all FDR < 1.05e^-10^) (***Figure 2c*; *Supplemental Table 4***). These observations demonstrate that the LIV-PM DE signatures identified here are reproducible in independent snRNA-seq data.

### Differing pathway annotations for cell-type-specific LIV-PM DE signatures

The large number of DEGs identified suggests that many biological processes may be affected by LIV-PM status. To verify this, biological processes defined in the Gene Ontology (GO) database^26^ were tested for enrichment of DEGs. The statistically significant GO gene set enrichment test results (FDR ≤ 0.05) for LIV DEGs and PM DEGs are provided in ***Supplemental Tables 5A and 5B.*** GO gene sets which show the most consistent (across the most cell types) and significant (lowest FDR) enrichment for LIV DEGs included terms related to energy metabolism, RNA processing, splicing, and myelination. Whereas, GO gene sets which show the most consistent and significant enrichment for PM DEGs included terms related to cell signaling, nervous system development, cellular response to stimuli, and protein modifications. While the LIV DEG and PM DEG associated GO enrichment presented here largely recapitulates what was presented in the Liharska *et al.*^23^, novel findings include the LIV DEG enrichment of transfer-RNA metabolism and processing in Oli nuclei, and the PM DEG enrichment of ubiquitin-dependent protein catabolic processes specific to Exc1 neurons. Additionally, the PM DEG enrichment of cell signaling previously identified in bulk RNA-seq extends across 8 of the 9 main cell types analyzed (***Figure 2d***). Together, the GO enrichment results presented here both replicates and extends the findings from Liharska *et al.*^23^ by providing a cell type level understanding of the biological processes associated with LIV-PM DEGs.

### Cell type marker genes are enriched for either LIV DEGs or PM DEGs

The results presented up until now suggest that cell type identity is conserved regardless of LIV-PM status but that within a cell type many genes are LIV-PM DEGs. These two observations led to the hypothesis that cell type markers would also be differentially expressed between LIV and PM samples (***Figure 2e*; *Supplemental Figure 4***). To test this hypothesis, for each of the 9 neural cell types, the cell type markers defined above were tested for enrichment of the corresponding cell type specific LIV or PM DEGs. Oli and MG cell type markers were significantly depleted of PM DEGs (Fisher’s exact tests ORs = 0.03 for Oli and 0.64 for MG; FDR < 0.04) and significantly enriched for LIV DEGs (ORs = 2.42 for Oli and 1.83 for MG, FDR < 3.14e^-4^). In contrast, Exc1, Exc3, Int2, and OPC cell type markers were significantly enriched for PM DEGs (Fisher’s exact tests ORs = 2.00 – 7.97, FDR = 1.46 x 10^-4^) and significantly depleted for LIV DEGs (ORs 0.00 – 0.43, FDR = 2.90 x 10^-3^). LIV DEGs were also found to be depleted of Int1 markers in Int1 nuclei (Fisher’s exact test OR = 0.00, FDR = 0.02) and enriched for Ast markers in Ast nuclei (Fisher’s exact test OR = 2.38, FDR = 1.07e^-6^). These findings show that while cell type identity is conserved regardless of LIV-PM status, the expression levels of the genes that define cell type identity are often associated with LIV-PM status. Moreover, these marker genes display opposite enrichments for LIV or PM DEGs, potentially impacting results from downstream analyses of snRNA-seq data that leverage cell type specific marker gene expression.

### Estimation of cell-type composition is significantly more accurate in living PFC mixtures

In a bulk RNA-seq experiment, the cell type composition of each sample is unknown, and analyses of this type of data may be confounded by sample differences in cell type compositions. At present, sample sizes of bulk RNA-seq datasets are generally orders of magnitude larger than sample sizes of snRNA-seq datasets. Therefore, one important use of snRNA-seq data is as a reference to estimate the cell type proportions of samples in bulk RNA-seq data (i.e., cell type deconvolution)^30^. Estimated cell type proportions are often used in statistical models to account for the unmeasured differences in cell type composition between samples. While previous work has shown that choice of reference or method used for cell type deconvolution influences the accuracy of cell type proportion estimates^27^, determining whether deconvolution accuracy is impacted by the LIV-PM status of either the snRNA-seq reference used or the bulk samples deconvolved (i.e., mixtures) has yet to be determined.

Given the availability of snRNA-seq data from both LIV samples and PM samples collected for this report, the following research questions were addressed: Is the accuracy of cell-type proportion estimates from cell type deconvolution affected by the LIV-PM status of the bulk RNA-seq mixtures being deconvolved, or the LIV-PM status of the snRNA-seq data used as reference. Using as input the 52 samples with snRNA-seq data generated for this report, the following procedure was performed: (1) a LIV reference dataset was created by randomly selecting 5 LIV samples, (2) a PM reference dataset was created by randomly selecting 5 PM samples; (3) mixtures to be deconvolved were generated as pseudo-bulk from all nuclei for each of the 42 remaining samples for which the true cell type proportions are known, and (4) the mixtures were deconvolved using the LIV and PM references. To account for the stochasticity introduced by the random selection of samples to serve as a reference, the procedure described above was performed 50 times. For each cell type, two estimates were calculated for each mixture: one from the LIV references (i.e., the mean of the estimates calculated using the LIV references) and one from the PM references (i.e., the mean of the estimates calculated using the PM references). The accuracy of these estimates was calculated as the absolute difference between the estimate and the true cell type proportion (i.e., the number of nuclei of that cell type / total number nuclei in the mixture). Regardless of the LIV-PM status of the samples used for the reference, deconvolution accuracy was significantly greater in LIV mixtures compared to PM mixtures for Exc1 (FDR: LIV reference = 5.29e^-3^, PM reference = 1.89e^-3^) and Ast (FDR: LIV reference = 1.72e^-5^, PM reference = 1.37e^-2^) (***Figure 3a***). When using the LIV reference, accuracy was significantly greater in LIV mixtures compared to PM mixtures for Int1 (FDR = 0.02) (***Figure 3a***). For PM mixtures, accuracy was significantly greater when (1) using PM references compared to LIV references for Exc3 in PM mixtures (FDR = 0.02) and (2) using LIV references compared to PM references for Int1 (FDR = 1.47e^-4^) (***Figure 3b***). No other comparisons were significant after adjusting for multiple testing. Together, these results suggest that, while the choice of LIV or PM references appear to have limited impact on deconvolution accuracy, expression differences between LIV and PM samples can result in higher accuracies of cell type estimations for a subset of neural cell types in LIV mixtures.

**Figure 3:**
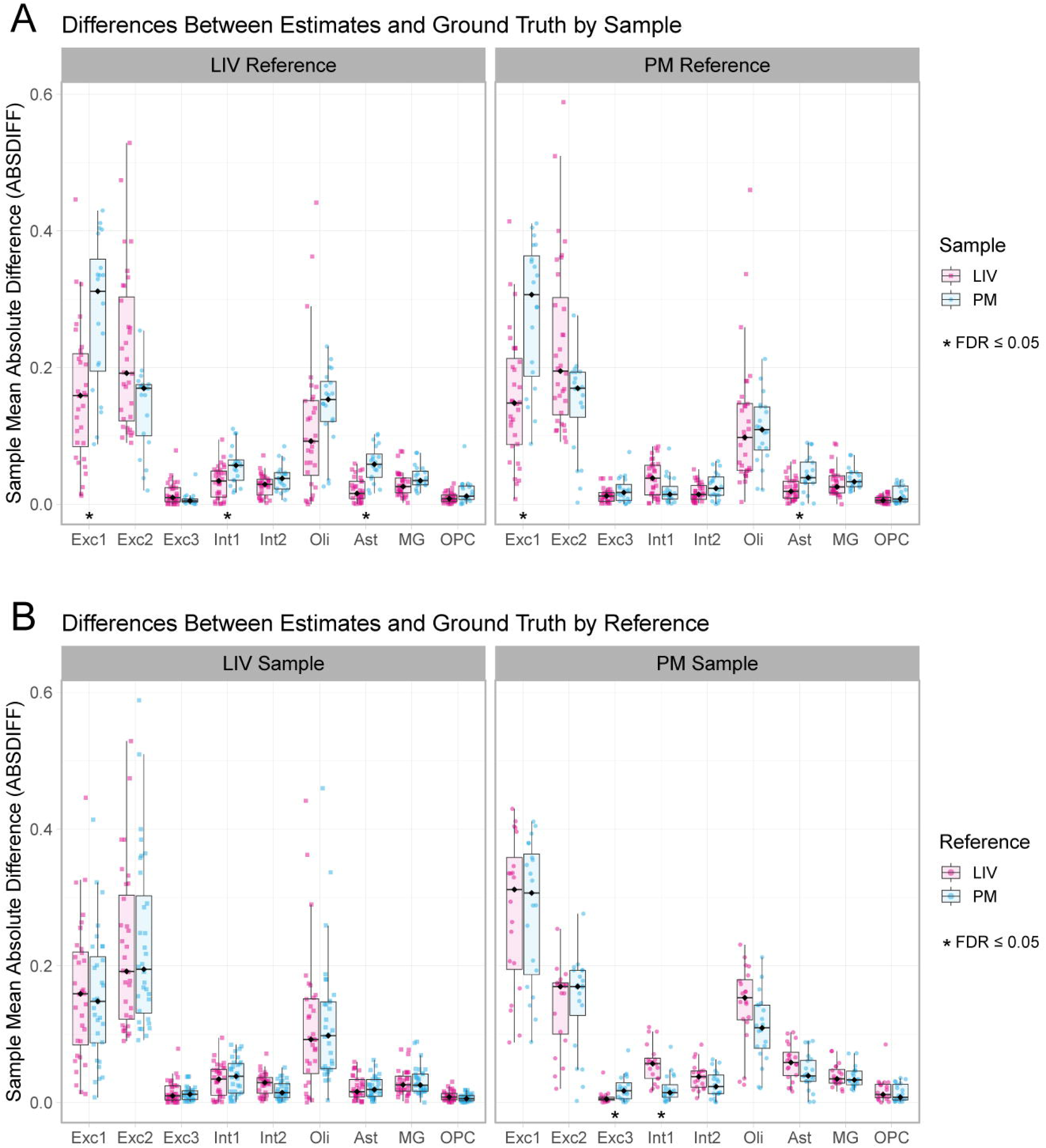
Assessing cell type deconvolution accuracy. The absolute value of the difference between CIBERSORTx cell type proportion estimates and true proportion (ABSDIFF; y-axis) for each cell type (x-axis). Each point and represents the average ABSDIFF per sample across all permutations of the deconvolution calculations, boxplots represent the median ABSDIFF per group (LIV or PM) depending on whether samples or references are being compared. Point shape represents the LIV-PM status of the mixtures being used in the comparison (LIV = square, PM = circle). Additionally, points and boxplots are colored based on the LIV-PM status of the sample (LIV = pink, PM = blue). ***A)*** ABSDIFF comparisons between estimates calculated on either LIV or PM samples faceted by whether a LIV or PM reference was used to estimate cell type proportions. Significant differences (*) between LIV and PM sample ABSDIFF is calculated based on Wilcoxon ranked-sum test and adjusted for multiple testing (FDR ≤ 0.05). ***B)*** ABSDIFF comparisons between estimates calculated from either LIV or PM snRNA-seq references faceted by whether predictions were performed on LIV or PM samples. Significant differences (*) in ABSDIFF between whether a LIV and PM reference was used is calculated based on Wilcoxon ranked-sum test and adjusted for multiple testing (FDR ≤ 0.05).

### Quantifying and accounting for differences between living and postmortem samples

One of the findings of Liharska *et al*^23^. was that disease-associated genes identified using RNA-seq in postmortem human brain tissues demonstrated a pattern of overlap with LIV-PM DEGs (i.e., case DEGs overlap PM DEGs and control DEGs overlap LIV DEGs), raising the possibility that brain gene expression signatures of neurological and mental illnesses may be confounded by differences observed between living and postmortem samples. To allow brain researchers to effectively model and account for LIV-PM differences in gene expression, an approach was developed to summarize the effects of LIV-PM status on the gene expression of a human brain sample into a single metric that can, in turn, be used to account for these effects.

Towards this end, the following procedure (***Figure 4a***) was performed 50 times on the pseudo-bulk data for each cell type from the 52 samples in the full snRNA-seq LBP cohort: (1) the 52 samples were randomly split into 3 sets (a training set, a testing set, and a holdout set) each comprised of both LIV samples and PM samples; (2) by applying elastic net binomial regression to the training set pseudo-bulk data, a model was derived to calculate the probability a set of gene expression values are from a postmortem sample (hereafter, this probability is referred to as “PMlink”); (3) the model was applied to the testing set pseudo-bulk data, resulting in a PMlink value for each sample in the testing set; (4) the ability of PMlink to classify a sample as either a LIV sample or a PM sample in the testing set was examined; (5) two versions of LIV-PM DE were performed on the testing set data – one version that included PMlink as a covariate in the model and one version that did not included PMlink as a covariate in the model – and the resulting LIV-PM DE signatures were compared to one another and the LIV-PM DE signature from the holdout set as reference; (6) DE of the PMlink variable was performed in only the PM samples in the testing set, resulting in a PMlink DE signature; (7) LIV-PM DE was performed in the samples of the holdout set, and the resulting LIV-PM DE signature was compared to the PMlink DE signature from the PM samples from the testing set. From this procedure, four observations were made. First, PMlink accurately classified LIV samples and PM samples in the testing set (receiver operating characteristic [ROC] - area under the curve [AUC] score ~ 1 for all permutations and cell types; ***Figure 4c*, *Supplemental Table 7***). Second, the median correlations between the two LIV-PM DE signatures identified in the testing set (i.e., the signature with PMlink as a covariate in the model (***Supplemental Table 8a***) and the signature without PMlink as a covariate in the model (***Supplemental Table 8b***) were 0.17 for Exc1, 0.35 for Exc2, 0.24 for Exc3, 0.12 for Int1, 0.23 for Int2, 0.20 for Oli, 0.36 for Ast, 0.35 for MG, and 0.30 for OPC, all of which were less than their respective reference LIV-PM DE comparison (***Figure 5a***). Third, including PMlink as a covariate in the LIV-PM DE analysis performed on the testing set decreased the number of LIV-PM DEGs identified by an average of 99% for Exc1, 50% for Exc2, 99% for Exc3, 90% for Int1, 99% for Int2, 92% for Oli, 98% for Ast, 54% for MG, and 96% for OPC (***Figure 5b***). Fourth, the PMlink DE signatures from the PM samples of the testing set (***Supplemental Table 8c***) were generally positively correlated with LIV-PM DE signatures from the holdout set (***Supplemental Table 8d***) (median ρ: Exc1 = 0.23, Exc2 = 0.07, Exc3 = 0.25, Int1 = 0.22, Int2 = 0.28, Oli = 0.25, Ast = 0.12, MG = 0.09, OPC = 0.21; ***Figure 5c***). The procedure described here for each cell type in the pseudo-bulk gene expression data was also performed on the all-nuclei pseudo-bulk and the bulk RNA-seq gene expression data and all of these findings were replicated (***Supplemental Figure 5***).

**Figure 4:**
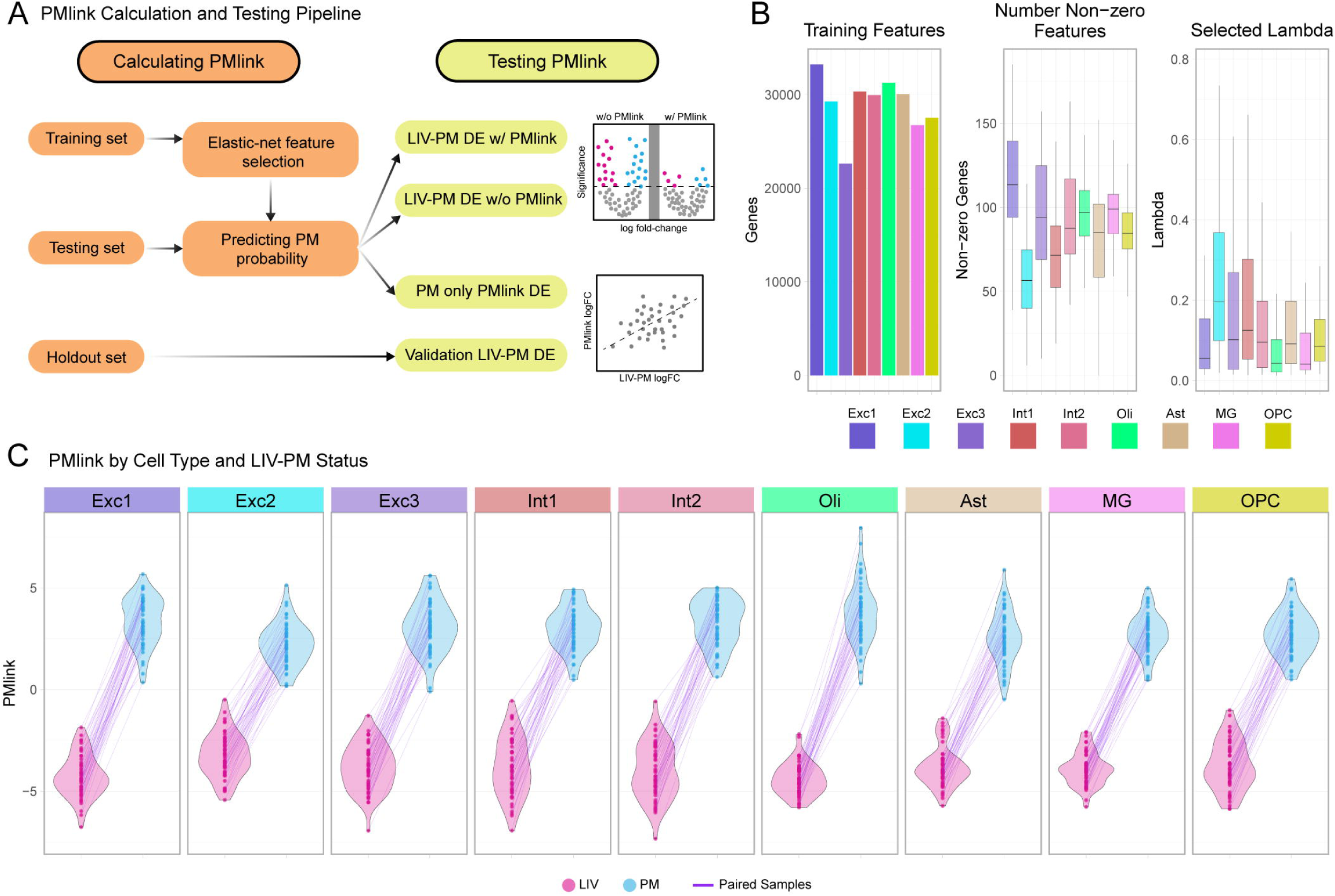
Calculating postmortem probability score (PMlink) across 50 testing, training, and holdout data splits. ***A)*** PMlink calculation and testing flowchart presenting how training, testing, and holdout sets are used in the calculation and validation of PMlink as a metric recapitulating expression difference between LIV and PM pseudo-bulk samples. ***B)*** Technical parameters (mean number of starting genes as features (left barplot), number of selected non-zero features after regularization (center boxplot), and the selected lambda used for postmortem probability predictions (right boxplot); y-axis) used for the 50 cross-validated elastic-net calculations colored by cell type. ***C)*** Violin plots presenting the distribution of PMlink scores (y-axis) calculated from the scaled postmortem probability across all 50 elastic-net iterations separated by the LIV-PM status of samples (LIV = pink, PM = blue). Each point represents a sample also colored by LIV-PM status with paired samples within the same testing set connected by a purple line. The violin plots are faceted for each of the 9 analyzed cell types for which PMlink scores were calculated.

**Figure 5:**
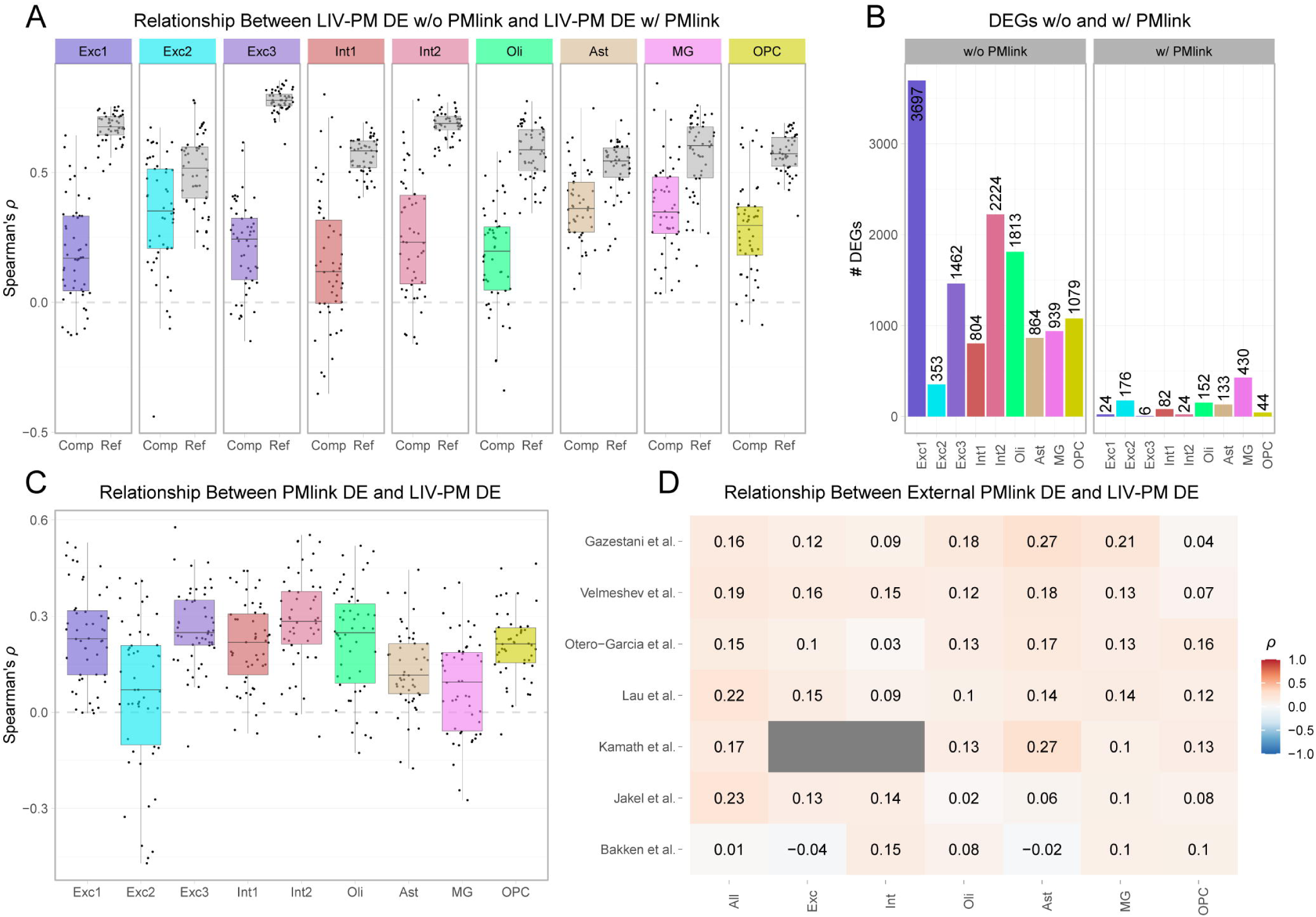
Testing the ability of PMlink to replicate LIV-PM DE across internal holdout sets and external snRNA-seq and bulk datasets. ***A)*** Spearman’s correlation coefficients (ρ; y-axis) comparing the logFCs from LIV-PM DE calculated on the testing set before and after including PMlink in the linear model. Points represent the ρ for each testing sample split iteration faceted by cell type in which PMlink is calculated. Within each facet, the colored boxplot represents the with and without PMlink comparison (Comp; x-axis) and the grey boxplot represents the reference comparison the testing set LIV-PM DE without PMlink and the holdout set LIV-PM DE (Ref; x-axis). ***B)*** Bar plots depicting the average number of significant DEGs (FDR ≤ 0.05) across testing set LIV-PM DE analyses before (left) and after (right) including PMlink in the simple linear model across all cell types (x-axis). ***C)*** Spearman’s correlation coefficients (ρ; y-axis) comparing the logFCs from LIV-PM DE calculated on the holdout set and PMlink DE calculated on the postmortem subset of each testing set. Points represent the ρ for each testing sample split iteration with the x-axis and colors representing the cell-type each comparison was performed in. ***D)*** Heat map of median Spearman’s ρ (low = blue, high = red) comparing logFCs from PMlink DE in the external snRNA-seq datasets (y-axis) with internal holdout set LIV-PM DE analysis for each of the shared annotated cell types across all datasets (x-axis).

Next, the ability of PMlink to identify the effects of LIV-PM status on gene expression was assessed in external human brain gene expression datasets^22,28–35^. External snRNA-seq datasets were obtained from a recently published repository of 16 postmortem snRNA-seq datasets and 1 snRNA-seq dataset obtained via neurosurgery^36^. From this repository, 6 snRNA-seq datasets (Bakken et al.^31^: samples=14 (PM), nulcei=74,813; Jäkel et al.^34^, samples=20 (PM), nuclei=10,783; Kamath et al.^32^: samples=18 (PM), nuclei=241,307; Lau et al..^35^: samples=21 (PM), nuclei=133,533; Otero-Garcia et al..^28^: samples=19 (PM), nuclei=50,478; Velmeshev et al..^33^: samples=41 (PM), nuclei=79,630; Gazestani et al..^36^: samples=52 (LIV), nuclei=892,828) were selected based on samples sizes (>10) and the number of annotated cell types present in the dataset (>4). For each of the external datasets studied, the following procedure was performed 50 times from cell type and all-nuclei pseudo-bulk for the 52 samples in the full snRNA-seq LBP cohort: (1) the 52 LBP samples were randomly split into 2 sets (a training set and a holdout set) each comprised of both LIV samples and PM samples; (2) the external dataset was defined as the testing set; (3) by applying elastic net linear regression to the training set pseudo-bulk data, a model was derived to calculate PMlink in the testing set; (4) the model was applied to the testing set cell-type and all-nuclei pseudo-bulk data, resulting in a PMlink value for each sample in the testing set; (5) DE of the PMlink variable was performed in the testing set, resulting in a PMlink DE signature; (6) LIV-PM DE was performed in the samples of the holdout set, and the resulting LIV-PM DE signature was compared to the PMlink DE signature from the testing set. The median of this correlation coefficient across 50 permutations of testing and holdout data splits was then averaged across the analyzed external datasets (ρ: Exc = 0.10, Int = 0.11, Oli = 0.11, Ast = 0.15, MG = 0.13, OPC = 0.10, All-nuclei = 0.16; ***Figure 5d***).

In addition to testing the ability of PMlink to capture LIV-PM expression differences in snRNA-seq data, we sought to assess whether PMlink trained and tested on bulk RNA-seq data could also recapitulate the correlations of LIV-PM and PMlink DE signatures observed above. The procedure described in the previous paragraph was thus repeated 50 times using: (1) the bulk RNA-seq presented in Lihaska *et al.*, splitting that data into training (26 samples) and holdout sets (26 samples); and (2) two external bulk RNA-seq datasets generated from the CommonMind Consortium research iniative as testing sets^30^ (CMC: n = 516; HBC: n = 305). Estimates of the correlation between LIV-PM and PMlink DE signatures were similar to those obtained when applying the PMlink procedure to the snRNA-seq data (median Spearman’s ρ: CMC= 0.16, HBC = 0.28, ***Supplemental Figure 5e***). Overall these results present and characterize a unique method to estimate and account for differences in gene expression between LIV and PM samples in both snRNA-seq and bulk RNA-seq datasets consisting exclusively of either PM samples or LIV samples.

## DISCUSSION

Understanding the molecular underpinnings of human brain function is a necessary step towards studying brain illnesses. Technological advances such as snRNA-seq have greatly improved the resolution of molecular signatures associated with disease, but molecular studies of human brain are limited by access to brain tissue. Although many important findings have emerged from the use of postmortem tissue^37–40^, results here and from the Lihaska *et al.*^23^ suggest that transcriptome-wide differences in gene expression between LIV and PM samples need to be recognized and contextualized for past and future postmortem brain studies. Analyses from a unique dataset containing snRNA-seq from both living and postmortem donors allows for the profiling of these expression differences across annotated neural cell types. Although snRNA-seq data from both LIV and PM nuclei generally clustered into the same neural cell types, DE analyses revealed consistent gene expression signatures across all nuclei and cell types, including for genes used to define many of these cell types. This DE signal was also shown to be present in a PD-only subset of the cohort, and replicated in both an independent bulk RNA-seq cohort^23^ and an external snRNA-seq dataset^12^. Key biological processes, such as neurosignalling, neurogenesis, stimulus response, and protein modifications, were identified as upregulated in PM samples while pathways such as energy metabolism, myelin synthesis, and RNA processing were found to be upregulated in LIV samples. Additionally, differences in gene expression between LIV and PM samples significantly impacted the accuracy of cell type deconvolution, with Exc1 and Ast cell type proportions more accurately estimated in LIV mixtures regardless of reference. Finally, we present a unique method (PMlink) that leverages data generated from this type of study to successfully identify and account for transcriptomic differences between LIV and PM samples in other datasets.

The PM DE signature identified by Liharska *et al.*^23^, which captures genes with higher expression in postmortem brain tissue than in brain tissue obtained through neurological procedure, and the biological processes associated with them, was characterized by an enrichment of stress response, apoptosis, and inflammation associated pathways^23^. This pattern of upregulation is consistent with changes that occur during periods of cerebral ischemia followed by hypoxia as the heart stops and blood circulation to the brain ceases^41–43^. The snRNA-seq PM DE signature presented here shows a subset of non-overlapping biological processes that may reflect cell type specific expression changes associated with perimortem ischemia. For example, increased neurogenesis and neurosignaling have been linked to cerebral ischemia as a neuroprotective factor to apoptosis^44,45^, and terms related to neurogenesis and neurosignaling appear amongst the most enriched pathways for PM DEGs across all cell types. One hypothesis for the increased neurosignaling reflected by the pathway enrichments for PM DEGs could be a response to neuronal depolarization in the low energy ischemic environment resulting in cytotoxic Ca+ concentrations and glutamate release, effectively triggering activity-dependent transcription factors in the nucleus ^46–48^. Additionally, in Exc1 PM DEGs we see enrichment of modification-dependent protein catabolism which represents an important mechanism for recycling biological material back into usable energy^49^, which may represent another protective response to the energy depleted conditions within dying neurons. Together, these findings suggest that genes with increased transcript abundance in PM samples may represent expression changes associated with biological processes responding to perimortem injury and pre-agonal ischemia/hypoxia in the dying brain.

As previously shown, the expression of genes involved in cellular respiration processes are detected at a higher level in living tissue^23^. This result is once again compatible with the presence of postmortem hypoxia/ischemia, as oxygen is no longer transported to the brain (>90% of the ATP present in the brain is depleted within 5 minutes)^50^. Here, genes with higher expression in LIV samples than in PM samples are enriched for many energy-dependent biological processes such as oligodendrocyte function and RNA metabolism. Oligodendrocytes represent the most energy demanding neural cell type and require high levels of ATP to perform their most basic function of myelin synthesis and maintenance^51^. This process has been shown to be especially susceptible to oxidative stress under pathological, hypoxic, and ischemic conditions^52,53^. Additionally, problems associated with myelination have been shown to have deleterious downstream effects which often results in pathology^54^. The vulnerability of oligodendrocytes to apoptosis in the postmortem environment is potentially captured by both the magnitude of the oligodendrocyte DE signal and the enrichment of myelination and oligodendrocyte development pathways in LIV DEGs across all cell types. Oligodendrocyte markers were also present at a significantly higher level in living nuclei across non-myelinating cells. This is consistent with previous reports showing myelin synthesis associated genes are expressed in the nuclei of neurons and other non-myelinating cells^55,56^. Beyond myelin associated genes, the other key process upregulated in LIV nuclei is RNA metabolism at various stages. Specifically, RNA splicing not only requires ATP hydrolysis for the RNA-protein rearrangements needed for spliceosome assembly, but also in the function of ATP-dependent RNA helicase, which helps regulate splicing^57^. Additionally, transfer-RNA (tRNA) metabolism shows significant enrichment for LIV DEGs, highlighting another energy dependent process important for initiating translation^58^. For tRNAs to interact with amino acids that will be incorporated into the polypeptide chain, they must first become charged via aminoacytlation, a process which requires ATP and is enriched among LIV DEGs across cell types but most significantly in oligodendrocytes^59^. The enrichment of energy-related processes in genes with increased abundance in living samples is reassuring based on the understanding that energy levels are depleted and cellular respiration is halted in the hypoxic perimortem and postmortem environment. More importantly, disruption of oligodendrocyte function and RNA transcription/translation emphasizes the need to understand the differences between LIV and PM samples to best interpret disease gene expression and protein signatures identified postmortem which implicate oligodendrocytes or transcriptional/translational regulatory factors in neuropathology.

The transcriptomic differences between living and postmortem brain tissue mentioned above present compelling evidence for the utility of living brain tissue samples as an additional resource in the study of human molecular neurobiology. As shown in the results section, clustering of cells in snRNA-seq data and the annotation of cell clusters to specific cell types was not impacted by the living/postmortem state of the tissue assessed. This was expected given the vast differences in gene expression that have been regularly observed between cell types, that often dwarfs those differences that could be explained by cell state (e.g., disease or, in this case living/postmortem states)^12,60,61^. This finding confirms that, to the best of our knowledge and for the purpose of generating brain cell type atlases or references, living and postmortem tissue can be interchangeably used, a crucial consideration given the relative complexity of living brain tissue access and the impossibility to obtain complete brains for molecular profiling from living individuals. However, the strength of the DE signal within these defined cell types has consequences for downstream applications of snRNA-seq data. Here, cell type deconvolution accuracy was assessed using independent LIV or PM references against LIV or PM pseudo-bulk mixtures to determine the impact of DE on this commonly used application of snRNA-seq data. Interestingly, while LIV-PM status of reference samples did not seem to impact the accuracy of the deconvolution procedure, for some cell types, the LIV-PM status of the mixtures showed significantly better accuracy for LIV mixtures. This could be explained by the choice of deconvolution method, where the one tested here (CIBERSORTx) uses a signature matrix, which is defined using cell type specific gene expression profiles from input sc/sn-RNAseq references^62^, to calculate estimates from bulk mixtures. Generating cell type specific signature matrices requires well defined marker genes^63^ and thus, DE of these markers might help explain differences in deconvolution accuracy between living and postmortem mixtures in our data. Living and postmortem references impact deconvolution accuracy in a meaningful way only for specific neuronal subtypes in PM mixtures, further highlighting that cell type identity of living and postmortem nuclei remains consistent and living snRNA-seq could be used as a suitable reference for deconvolution procedures, especially for broad neuronal and glial cell types.

This study also provides and characterizes an elastic-net based approach that was employed to successfully and effectively account for the differences in gene expression between LIV and PM samples when included as a covariate in the linear mixed model used for differential expression purposes. The development of this method that, using similar underlying concept as deconvolution, identifies and accounts for differences in gene expression observed between LIV and PM samples has the potential to help brain molecular researchers to better understand healthy and diseased brain neurobiology. This approach is also analogous to disease risk prediction methods, in which computationally selected factors (genes, genetic variants, phenotypic variables, etc.) are used to generate scores that represent risk approximations^64–66^. The transferability of this method was also tested through replicating LIV-PM DE using PMlink calculated on external snRNA-seq datasets curated as part of another study^22^ and selected bulk RNAseq datasets^29,30^. The success of this preliminary assessment of a method to correct for gene expression differences between LIV and PM samples reflects the strength of the overall DE signal between LIV and PM samples which allowed for near perfect accuracy when predicting LIV or PM state in independent pseudo-bulk and bulk testing datasets. Additional investigation is still required to better understand how study design and cell type specific variation impacts the ability for PMlink to recapitulate our LIV-PM DE results. It is important to note that the preliminary implementation of this correction method likely represents the lower bounds for a meaningful postmortem linear predictor variable such as PMlink. Furthermore, to avoid data leakage, LBP datasets needed to be split into testing, training, and holdout sets, greatly reducing the statistical power for PMlink and LIV-PM DE comparisons, and likely contributing to the variance among correlation coefficients. A curated training set of samples and/or nuclei that reflect LIV-PM differences with low variability across other dimensions might improve elastic-net accuracy and consistency in generating postmortem probabilities. New large-scale snRNA-seq atlases of the human postmortem brain^67^ provide robust resources for developing, optimizing, and testing a scalable correction method to account for living and postmortem expression differences in future studies of human neurobiology and brain disease.

The novel characterization of living and postmortem expression difference within in the context of common snRNA-seq approaches presented in this report has a number of limitations and remaining questions that will require further investigation. Most prominent is the inability to, given the study design, account for all possible confounder of gene expression, such as those that might be caused by either the neurosurgical procedure or any other technical effect unique to LIV samples. While results were, when possible, replicated using many independent resources, functional validation studies in animal or cell models were not performed as part of this study. Such experimental studies may, beyond simply validating the LIV-PM differences observed here, help assess the putative confounding effects described above in a highly controlled setting. Additionally, as it is not possible to obtain brain tissue from healthy individuals neurosurgically, the living cohort lacks neurotypical controls and is limited to mainly PD diagnosed individuals. Although LIV-PM DE performed on only PD individuals almost perfectly recapitulates the original LIV-PM DE signal, future studies should focus on corroborating these results using different clinical phenotypes and neurosurgeries. Expanding biopsy procedures to different neurosurgeries will also provide the opportunity to profile LIV-PM expression differences in brain regions other than the PFC. Finally, while we have shown that there are vast expression differences between living and postmortem samples that need to be taken into consideration for future snRNA-seq studies of the human brain, it is important to note that this alone does not prove that either living or postmortem tissue is superior to the other for profiling disease signatures in the human brain.

Overall, this study provides the first characterization of: (1) snRNA-seq expression differences between LIV and PM brain samples, (2) the potential biological underpinnings of this DE signal, and (3) a novel approach to integrate living samples into future postmortem snRNA-seq study designs to account for differences in gene expression observed between LIV and PM samples. Beyond results presented here, the use of living tissue that was ethically biopsied at scale with no recorded deleterious consequences to patients for the sole purpose of research presents an exciting future for the study of the human brain. Finally, this study presents a large body of evidence that many gene expression differences exist at the single nuclei level between LIV and PM samples, highlights the utility of living brain tissue as an emerging resource to study the brain, and suggests that the use of postmortem tissue for the study of human molecular neurobiology could benefit from being integrated with knowledge derived from living brain tissue.

## METHODS

### Ethics Statement

All living cohort participants consented to sample collection, genomic profiling, clinical data extraction from medical records, and public sharing of de-identified data as part of STUDY-13-00415 of the Human Research Protection Program at the Icahn School of Medicine at Mount Sinai. All IDs for living tissue samples have been de-identified and are not traceable back to patients.

### Living and Postmortem Cortical Samples

Living (LIV) PFC samples were collected from patients undergoing deep-brain stimulation surgery. For a detailed description of the living cohort, the adapted neurosurgery protocol, and post biopsy tissue processing refer to the Liharska *et al*^23^. Postmortem (PM) PFC samples used for this study were obtained from two different postmortem brain banks: Harvard Brain Tissue Resource Center and New York Brain Bank at Columbia University. PM samples were matched to LIV samples in respect to age and sex when possible. The standard PM tissue processing following autopsy was performed as described previously^68,69^,. Lab protocols for storing postmortem tissue samples are also described in Liharska *et al*^23^. In total, 9 unilateral biopsies and 13 bilateral biopsies were collected from 23 LIV individuals (LIV samples= 35) and 23 PM samples were selected for single-nuclei RNA isolation and processing.

### Nuclei Extraction with Kit

Nuclei were extracted from 58 frozen brain tissue samples (35 LIV and 23 PM) using the Minute Single Nucleus Isolation Kit for Neuronal Tissues and Cells (Invent Biotechnologies, #BN-020) following the manufacturer’s protocol. Briefly, around 10-20mg of frozen tissue was added to an Eppendorf tube. 200uL of cold buffer was added in with 5uL of RNase inhibitor and homogenized. Another 500uL of cold buffer was added to the tube with 12.5uL of RNase Inhibitor and further homogenized. The sample was then incubated on ice for 5 minutes (min). After incubation, the cell lysate was pipetted through a filter and into a collection tube. The sample was then incubated at –20C for 10 min. After incubation, the sample was centrifuged at 13000xg for 20 seconds (sec). Next, the filter was discarded and the pellet was suspended via pipette followed by another centrifuge step (600xg for 5 min). From this step, the supernatant was discarded, and the pellet was resuspended in 200uL 5% BSA in PBS with 5uL of RNase inhibitor. Next, 25uL of RNase inhibitor was added with another buffer into a new Eppendorf tube with 200uL of nuclear suspension fluid. This tube was then centrifuged at 1000xg for 10 min and the milky layer was then carefully removed via pipette. The rest of the supernatant was then removed without disturbing the pellet, and the pellet was resuspended in 200uL of 1x PBS. Finally, the quality and quantity of the extracted nuclei was checked with a cell counter.

### 10x Chromium Single Cell 3’ Gene Expression Profiling

After single nuclei isolation, 58 samples were processed with the 10x Chromium Single Cell Gene Expression assay using the Chromium Next GEM Single Cell 3′ Reagent Kits v3.1 (10X Genomics, #CG000204 Rev D). Gel bead emulsion (GEM) barcoding and subsequent library generation was performed following the manufacture’s protocol. Briefly, a target of 700-1200 cells/uL were loaded into a chip and then loaded into the Chromium machine. GEM quality was visibly checked to ensure that no clogs occurred before GEMs proceeded to the cleanup stage. After cleanup, cDNA was generated through PCR amplification. The quality of the cDNA was checked on an Agilent TapeStation using the High Sensitivity DNA ScreenTape Analysis (Agilent, #5067-5592) and quantification of cDNA was determined using the Qubit dsDNA HS Assay Kit (ThermoFisher, #Q32851). Following cDNA amplification, 3′ gene expression libraries were constructed using 10uL of cDNA product. Next, the number of total amplification cycles were determined based on the quantity of cDNA input as part of the sample index PCR step following the chart provided in the manual. After the libraries were constructed, quality was checked using the Agilent TapeStation and quantity checked with the Qubit dsDNA HS Assay Kit. 3 samples (2 LIV and 1 PM) failed QC during GEM preparation step stage and were not included for sequencing.

### Single Cell Sequencing of 3′ Gene Expression Libraries

After library construction, the libraries were pooled for sequencing. Before sequencing the pooled libraries were again checked for quality and quantity with the Agilent TapeStation. Sequencing was done on the NovaSeq with the NovaSeq 6000 S2 Reagent Kit v1.5 kit (Illumina, #20028315) for a target of 300M reads per sample. Sequencing followed the suggested 10X Chromium protocol generating reads with 28 base pairs for the cell barcodes and UMI, reads with 8 base pairs for the index sequence, and reads with 91 base pairs for the main inserts used for transcript quantification.

### Processing Raw Sequencing Reads and Sample Level QC

Raw fastq files were generated from sequenced libraries for 55 samples (33 LIV and 22 PM). *CellRanger* software (pipeline ver. 7.0) was used for genome alignment against the GRCh38-2020-A reference, generating BAM files and raw count feature-barcode matrices representing reads (UMI counts) aligned to genes (features) for each barcode assigned to a droplet. Reads mapped to introns and exons were incorporated into final count matrices in order to include both pre-mRNA and mature mRNA, which is representative of nuclear RNA populations. 1 LIV sample was removed for downstream analyses based on low number of cells relative to the rest of the samples. Identity concordance of snRNA-seq, bulk RNAseq, and whole genome sequencing (WGS) data is described in Liharska *et al*^23^. 1 PM sample was removed after identifying potential discrepancies between WGS and snRNA-seq data.

### Cell Level QC

Raw UMI feature-barcode count matrices which contains all droplets with valid GEM barcodes were first filtered in *CellRanger* (pipeline ver. 7.0) by removing “empty droplets” defined using the *EmptyDrops* procedure^70^. Briefly, empty droplets are defined from raw count matrices by first classifying high RNA count droplets as nuclei, and then calculating the cell-free expression profile of the remaining low count droplets, presumed to represent the ambient RNA in the fluid surrounding nuclei from destroyed or lysed cells. Barcodes of droplets containing expression profiles similar to the cell-free expression profile are then removed from the raw count matrices to generate filtered UMI count matrices for downstream processing. Removal of contaminating ambient RNA from nuclei containing droplets was done via SoupX (ver. 1.3.6), which takes the cell-free expression profile and background-corrects the filtered UMI count matrices accordingly^71^. Background-corrected count matrices were loaded into R (ver. 4.1) via Seurat (ver. 3.0)^72^. Empty droplets not previously identified and low quality cells were removed if the total number of unique genes per droplet and UMI counts per droplet was less than 200 or if greater than 1% of counts originated from mitochondria RNA (mt%). This conservative mt% threshold was selected for QC to reflect that snRNA-seq data should contain close to 0% mitochondrial RNA under ideal circumstances^73^. 1 LIV sample was removed for containing a high mt% in a majority of cells. Droplets predicted to contain multiple nuclei, termed doublets, were classified and removed using *scDblFinder*, a simulation and cluster based doublet detection method. The *scDblFinder* package (ver. 3.16) calculates a score for each cell based on the predicted expression profile of simulated doublets within a single sample^74^. To classify cells for doublet removal, count matrices from all samples were merged, normalized and clustered as described below, then an outlier-based threshold was used to remove clusters with a higher mean doublet score than average calculated doublet score. Then, all droplets not removed from outlier clusters with a doublet score > 0.25 were removed. This is a conservative threshold relative to the recommended threshold of 0.5^74^.

### Normalization, Integration and Clustering

For each sample (n = 52, LIV = 31, PM = 21), filtered and background-corrected UMI count matrices containing nuclei (droplets that passed cell level QC) were merged together and normalized using a scale factor of 10,000. The top 2000 highly variable genes were selected for principal component analysis (PCA) using the “vst” method within the *Seurat* package. From the first 15 principal components, *Harmony* (ver. 1.1)^75^ was utilized to perform a sample level correction to remove any variability that could confound cell type determination in dimensionally reduced space. To visualize the Harmony-corrected PCA embedding, the uniform manifold approximation and projection (UMAP) coordinates were calculated via the RunUMAP() function within *Seurat*. Next, the FindClusters() function was used for cluster identification, which performs Louvain-algorithm-based shared nearest neighbor (SNN) modularity optimization based clustering by calculating the k-nearest neighbors from Harmony embedding followed by the construction of an SNN graph^72^. A low clustering resolution of 0.6 was selected to isolate larger clusters that could be annotated into broad cell types. This normalization and integration process was performed prior to doublet removal, to identify and remove outlier doublet clusters, and then again on the final nuclei count after low quality droplets and doublets were removed.

### Cell Type Annotation

SNN clustering revealed 23 individual clusters from the integrated dataset. In an effort to annotate neural cell types from these 23 clusters, a marker gene analysis via the FindAllMarkers() function in *Seurat* was performed. Markers are defined as genes significantly upregulated (FDR ≤ 0.05) in one cluster relative to all other clusters as calculated using a Wilcoxon ranked sum test^72^. Markers calculated here were then cross-referenced to other snRNA-seq studies and literature on canonical markers for neuronal and glial cell types to merge and classify similar smaller clusters into 10 broad neural cell types representing excitatory neurons (Exc1, Exc2, and Exc3), inhibitory neurons (Int1 and Int2), oligodendrocytes (Oli), astrocytes (Ast), microglia (MG), oligodendrocyte progenitor cells (OPC), and non-neural cells (NonNeu). Non-neural cells represent endothelial or epithelial cells and were omitted from all downstream analyses.

### Scoring LIV and PM Marker Gene-sets

After annotation, samples were split into LIV and PM subsets and normalized, *Harmony* corrected, and clustered using the same parameters defined in the “Normalization, Integration and Clustering” section above. Marker genes from the original cell type annotations (Exc1, Exc2, Exc3, Int1, Int2, Oli, Ast, MG, OPC, and NonNeu) were then defined for the LIV subset and PM subsets independently using the procedure described in “Cell Type Annotation”. Next, a score representative of cell type identity was calculated for each nucleus from the aggregate expression of each marker gene set calculated in LIV samples and marker gene sets calculated in PM samples. This was completed for each nucleus using the AddModuleScore() function, which calculates the average expressions of selected gene sets subtracted by the aggregated expression of control genes selected randomly from all genes with a similar distribution of expression to the gene set in question^76^. The calculated scores for each cell type for either LIV or PM defined marker set were then compared across all nuclei using a Spearman’s correlation coefficient (ρ).

### Generating Pseudo-bulk

For downstream analyses (differential expression, cell type deconvolution, and PMlink analyses) nucleus-level counts were converted into pseudo-bulk counts representing gene expression at the sample level by aggregating counts per sample. Pseudo-bulk was generated as the sum of counts per sample within each cell type (i.e. Exc1 pseudo-bulk, Exc2 pseudo-bulk, etc.) and across all-nuclei for a given sample (all-nuclei pseudo-bulk) using the aggregateAcrossCells() function within the *scuttle* package (ver. 1.12)^77^.

### Differential Expression Analysis

To assess differential expression between living and postmortem nuclei, raw pseudo-bulk counts are first filtered to remove genes with low expression (sum of all counts per across all samples in either the LIV or PM group <10 for cell specific pseudo-bulk and <20 for all-nuclei pseudo-bulk). Normalization factors were calculated based on library size using the *calcNormFactors()* function in edgeR (ver. 3.14)^78^ and counts were transformed to log2-counts per million (logCPM) with observation-level weights based on the mean-variance relationship^72^. The relevant demographic covariates included as random effects in the linear mixed model (LMM) were sex, PD phenotype, and batch. Then, based on the correlation of technical covariates to the first 5 principal components following a principal component analysis of the normalized data, the final formula used for the LMM defining all LIV-PM differential expression analyses was:

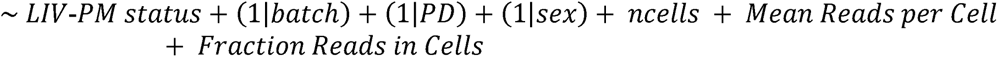

where the syntax “(1|V)” denotes a random effect for categorical variable V, while all other variables are modeled as fixed effects. Differential expression (DE) with LIV-PM status as the dependent variable was then calculated using the *dream* function from the *variancePartition* package (ver. 1.29)^79^, which is designed to use LMMs in calculating DE from *voomWithDreamWeights* normalized count matrices^80^. DE was calculated for each cell type pseudo-bulk count matrix and the all-nuclei pseudo-bulk count matrix. Significant differentially expressed genes (DEGs) were determined based on a FDR ≤ 0.05 after using the Benjamin-Hochberg method for multiple testing correction^81^. Each list of DEGs was further subdivided into LIV DEGs as the genes with higher abundance in living samples, denoted by with negative log fold-changes (logFCs); and PM DEGs as the genes with higher abundance in postmortem samples, denoted by positive logFCs.

### Replication of Differential Expression

Replication of all-cells pseudo-bulk DE was performed in an external bulk RNAseq dataset generated from living and postmortem cortical tissue as part of another LBP report (n=499, LIV=275, PM=224)^23^. Two changes were made from the original study when utilizing this dataset for the purpose of replication: 1) Any overlapping samples between the cohorts were removed from this replication cohort, and 2) neuronal cell fraction was omitted as a covariate in the LMM to maintain DE that might be specific to unique cell types. Spearman’s correlation was used to examine the relationship between the discovery (pseudo-bulk) and replication (bulk) logFC Next, LIV-PM DE was calculated in an independent single-nuclei dataset additional replication^12^. Since a majority of the Hodge *et al.* dataset represents NeuN+ sorted neurons, clusters representative of these neurons were identified by examining the expression of RBFOX3, the gene that encodes the NeuN protein. Only DE signatures from these clusters expressing RBFOX3 in at least 50% of cells were compared to the LIV-PM DE signatures define in Hodge *et al.*^12^. Next, the overlap between the LIV DEGs or PM DEGs identified here was compared to the 695 DEGs identified in the Hodge et al paper which contrasts nuclear gene expression of middle temporal gyrus (MTG) samples obtained from four living individuals to neuronal gene expression in the MTG of four postmortem donors. Enrichment tests were performed via a one-sided Fisher’s exact test to quantify overlap between (1) LIV DEGs across RBFOX3 expressing cell types from the LBP dataset and Hodge *et al.* LIV DEGs, and (2) PM DEGs across RBFOX3 expressing cell types from the LBP dataset and Hodge *et al.* PM DEGs. Odds ratios were calculated from these Fisher’s exact tests and significance was determined after adjusting for multiple testing (FDR ≤ 0.05). To test the impact of Parkinson’s disease on the LIV-PM DE signal across cell types, the same DE pipeline was utilized in only the PD diagnosed samples. Once again, all DE comparisons to test reproducibility of the LIV-PM signature were done using a Spearman’s correlation of logFC from the original LIV-PM DE and logFC from the replication LIV-PM DE.

### Biological Process Gene Ontology Enrichment

Biological processes gene ontology (GO) enrichment was performed from the full biological processes ontology term set^82^ using *topGO* (ver. 3.16)^83^ on the LIV DEGs and PM DEGs from the primary DE analyses on pseudo-bulk generated for the 9 main cell-types and the all-nuclei set. Significant GO term enrichment was determined using a Fisher’s exact test (FDR ≤ 0.05) with all genes defined as expressed (sum of all counts per across all samples in either the LIV or PM group <10 for cell specific pseudo-bulk and <20 for all-nuclei pseudo-bulk) for the background. Summarized GO term lists were generated from annotated terms containing <2500 genes, as to remove non-informative broad GO terms.

### Cell Type Deconvolution

Pseudo-bulk mixtures were deconvoluted via CIBERSORTx^62^ using 50 permutations of non-overlapping LIV or PM references. First, 50 subsets of 10 random samples (5 living and 5 postmortem) each were selected to be used as the LIV and PM single-cell references respectively. Each reference set was used for deconvolution with the existing 42 non-overlapping pseudo-bulk mixtures. Then, for each reference defined as described above iterations of CIBERSORTx were run with the standard recommended settings^62^ to generate predicted cell type proportions for each sample (100 CIBERSORTx total runs generating ~100 sets of estimated cell type fractions per mixture). The predicted cell type proportions were averaged across all permutations to get a single LIV and PM reference prediction for each mixture. The accuracy of the predicted deconvolution estimates was quantified by calculating the absolute value of the differences between mean estimate per mixture and that same mixture true cell proportions (sum of nuclei in a specific cell type / total nuclei per sample). Finally, significant differences between estimates calculated on LIV or PM mixtures and estimates calculated from LIV or PM references were tested via Wilcoxon ranked sum test and corrected for multiple testing using the Benjamini-Hochberg procedure.

### Calculating a Postmortem Prediction Score via ElasticNet

To create a score capable of accounting for gene expression difference between LIV and PM samples, a single continuous variable capturing the probability for a given sample to be classified as postmortem (PMlink) was calculated using a cross-validated elastic-net binomial regression via cv.glmnet() (ver.4.1). First, cell level and all-nuclei pseudo-bulk were split into three groups of approximately equal sizes: training (samples for elastic net feature selection; LIV = 10±1, PM=7±1), testing (sample to be assigned a PMlink score, LIV=10±1, PM=7±1), and holdout (holdout samples for LIV-PM DE; LIV = 11±1, PM=7±1). For PMlink calculations, the training set and testing set are first subset to include only genes with > 0 counts in at least one sample. Then the training set and testing set are normalized independently using *voom* from the *limma* package (ver. 3.58)^84^ and additional genes were removed so only genes present in both datasets were retained. The normalized training dataset was scaled and centered to 0 and the normalized testing dataset was scaled to the mean and standard deviation of the scaled training set. Using the normalized and scaled training set with genes as features, the elastic net binomial regression performs a combination of L1 (LASSO, α = 1) and L2 (ridge, α = 0) regularization using an α = 0.5 to shrink irrelevant feature coefficients to 0. The optimal regularization penalty (λ) is calculated through 10-fold cross-validation. We choose the largest value of λ (i.e. the most regularized model) for which cross-validated error is the smallest (lambda.min)^85^. Using the fitted binomial elastic-net model of non-zero coefficients in concert with the optimized λ, PMlink was calculated as the scaled probability of a sample from the normalized and scaled testing set being a postmortem sample. This process was then repeated with each pseudo-bulk sample being randomly assigned to either the training, testing or holdout set 50 times. PMlink calculations were also done on the external bulk data generated following the same procedure for the initial LBP flagship report^23^.

### Validating PMlink

We first assessed the accuracy of each PMlink score in classifying a sample’s origin as postmortem or living within each of the 50 permutations by calculating the area under the ROC curve (AUC) via the prediction() and performance() function from the *ROCR* package (ver. 1.11)^86^. The AUC represents the degree of separation between linear predictor scores with respect to actual LV-PM status of a sample by comparing false positives (FP) and true positives (TP). We then validated the utility of PMlink as a continuous metric for recapitulating the LIV-PM DE signature in two ways. (1) We assessed changes to the LIV-PM DE signal calculated from each testing set when PMlink is added as a covariate in a linear model with LIV-PM status as the dependent variable (DV). From the resulting two DE analyses per random split, we compared the number of number of DEGs (FDR ≥ 0.05) with and without including PMlink as an additional fixed effect in the LIV-PM DE model. Overall changes to the LIV-PM DE signatures were assessed by comparing logFCs from each DE analysis (with PMlink and without PMlink) using Spearman’s correlation (ρ). These correlations coefficients were then compared to a reference of Spearman’s correlation coefficients calculated between logFCs from LIV-PM DE calculated on the testing set without PMlink in the model and LIV-PM DE logFCs calculated on the holdout set. (2) We performed a DE analysis on only the PM samples from each testing set substituting PMlink as the primary DV in a linear model (no additional covariates); performed a second DE analysis for comparison on the holdout set using LIV-PM status as the primary DV in a simple linear model; and compared the logFCs from these 2 analyses using a Spearman’s correlation (ρ).

### Selecting and Processing External Datasets of PMlink Replication

Publicly available external datasets were selected to test if the PMlink calculation procedure presented here is informative to other snRNA-seq and bulk gene expression studies of human brain. To select adequate snRNA-seq datasets for testing, we focused on datasets that were curated as part of a recent study^36^ in which 26 human and mouse single-cell and single-nuclei RNA-seq datasets were integrated together and then re-annotated to broad neural cell types (excitatory neurons, inhibitory neurons, oligodendrocytes, astrocytes, microglia, oligodendrocytes progenitor cells, and endothelial). From this repository, 6 snRNA-seq datasets^28,31–36^ were chosen based on: (1) number of unique samples > 10 and (2) cell types overlapping with the LBP snRNA-seq dataset > 4. In addition to testing PMlink on external snRNA-seq datasets, we selected two bulk RNA-seq datasets generated as part of the CommonMind Consortium to assess the informativeness of PMlink derived from the bulk data presented in Liharska *et al*^23,30^. PMlink calculations on the external datasets were performed using the approach mentioned above with the following exception: the pseudo-bulk generated from the LBP snRNA-seq dataset and the bulk RNA-seq data presented in Liharska *et al.*^23^ were split evenly between training and holdout 50 times. The full external snRNA-seq pseudo-bulk and bulk datasets were used as the testing dataset. To test the ability of PMlink to recapitulate the LIV-PM DE signature presented here, two DE analyses were performed: (1) DE performed in the external pseudo-bulk and bulk datasets with PMlink as the DV in simple linear model and (2) LIV-PM DE calculated in the holdout pseudo-bulk. The relationship between these two DE analyses was assessed through comparing logFCs using a Spearman’s correlation (ρ).

## DATA AND MATERIALS AVAILABILITY

All consented data for the Living Brain Project, including, but is not limited, to raw snRNA-seq data and Bulk RNA-seq data used for this study is available on Synapse (https://www.synapse.org - Project SynID: syn2580853). Upon publication in a peer-reviewed journal, code for reproducing results from deposited data will be provided on GitHub.

## ACKNOWLEDGMENTS

The Living Brain Project study participants are commended for their important role in science. The following centers, programs, departments and institutes within the Icahn School of Medicine at Mount Sinai supported the work: Department of Neurosurgery; Department of Genetics and Genomic Sciences; Department of Psychiatry; Department of Neuroscience; Department of Medicine; Friedman Brain Institute; Department of Anesthesia; Department of Pathology; Charles Bronfman Institute for Personalized Medicine; Blau Center. Postmortem samples from Harvard Brain Tissue Resource Center were acquired under the National Institutes of Health NeuroBioBank request number 543. Postmortem samples from the New York Brain Bank of Columbia University were acquired under request number 1962.

## REFERENCES

1. Visscher, P.M., Wray, N.R., Zhang, Q., Sklar, P., McCarthy, M.I., Brown, M.A., and Yang, J. (2017). 10 Years of GWAS Discovery: Biology, Function, and Translation. The American Journal of Human Genetics 101, 5–22. 10.1016/j.ajhg.2017.06.005.

2. Lake, B.B., Ai, R., Kaeser, G.E., Salathia, N.S., Yung, Y.C., Liu, R., Wildberg, A., Gao, D., Fung, H.-L., Chen, S., et al. (2016). Neuronal subtypes and diversity revealed by single-nucleus RNA sequencing of the human brain. Science 352, 1586–1590. 10.1126/science.aaf1204.

3. Kotliar, D., Veres, A., Nagy, M.A., Tabrizi, S., Hodis, E., Melton, D.A., and Sabeti, P.C. (2019). Identifying gene expression programs of cell-type identity and cellular activity with single-cell RNA-Seq. eLife 8, e43803. 10.7554/eLife.43803.

4. Al-Dalahmah, O., Sosunov, A.A., Shaik, A., Ofori, K., Liu, Y., Vonsattel, J.P., Adorjan, I., Menon, V., and Goldman, J.E. (2020). Single-nucleus RNA-seq identifies Huntington disease astrocyte states. Acta Neuropathologica Communications 8, 19. 10.1186/s40478-020-0880-6.

5. Jiang, J., Wang, C., Qi, R., Fu, H., and Ma, Q. (2020). scREAD: A Single-Cell RNA-Seq Database for Alzheimer’s Disease. iScience 23, 101769. 10.1016/j.isci.2020.101769.

6. Nagy, C., Maitra, M., Tanti, A., Suderman, M., Théroux, J.-F., Davoli, M.A., Perlman, K., Yerko, V., Wang, Y.C., Tripathy, S.J., et al. (2020). Single-nucleus transcriptomics of the prefrontal cortex in major depressive disorder implicates oligodendrocyte precursor cells and excitatory neurons. Nat Neurosci 23, 771–781. 10.1038/s41593-020-0621-y.

7. Ahmadi, A., Gispert, J.D., Navarro, A., Vilor-Tejedor, N., and Sadeghi, I. (2021). Single-cell Transcriptional Changes in Neurodegenerative Diseases. Neuroscience 479, 192–205. 10.1016/j.neuroscience.2021.10.025.

8. Avila Cobos, F., Alquicira-Hernandez, J., Powell, J.E., Mestdagh, P., and De Preter, K. (2020). Benchmarking of cell type deconvolution pipelines for transcriptomics data. Nat Commun 11, 5650. 10.1038/s41467-020-19015-1.

9. Wang, X., Park, J., Susztak, K., Zhang, N.R., and Li, M. (2019). Bulk tissue cell type deconvolution with multi-subject single-cell expression reference. Nat Commun 10, 380. 10.1038/s41467-018-08023-x.

10. Stark, R., Grzelak, M., and Hadfield, J. (2019). RNA sequencing: the teenage years. Nat Rev Genet 20, 631–656. 10.1038/s41576-019-0150-2.

11. Tasic, B., Yao, Z., Graybuck, L.T., Smith, K.A., Nguyen, T.N., Bertagnolli, D., Goldy, J., Garren, E., Economo, M.N., Viswanathan, S., et al. (2018). Shared and distinct transcriptomic cell types across neocortical areas. Nature 563, 72–78. 10.1038/s41586-018-0654-5.

12. Hodge, R.D., Bakken, T.E., Miller, J.A., Smith, K.A., Barkan, E.R., Graybuck, L.T., Close, J.L., Long, B., Johansen, N., Penn, O., et al. (2019). Conserved cell types with divergent features in human versus mouse cortex. Nature 573, 61–68. 10.1038/s41586-019-1506-7.

13. DeMaio, A., Mehrotra, S., Sambamurti, K., and Husain, S. (2022). The role of the adaptive immune system and T cell dysfunction in neurodegenerative diseases. J Neuroinflammation 19, 251. 10.1186/s12974-022-02605-9.

14. Kamath, T., Abdulraouf, A., Burris, S.J., Langlieb, J., Gazestani, V., Nadaf, N.M., Balderrama, K., Vanderburg, C., and Macosko, E.Z. (2022). Single-cell genomic profiling of human dopamine neurons identifies a population that selectively degenerates in Parkinson’s disease. Nat Neurosci 25, 588–595. 10.1038/s41593-022-01061-1.

15. Fullard, J.F., Lee, H.-C., Voloudakis, G., Suo, S., Javidfar, B., Shao, Z., Peter, C., Zhang, W., Jiang, S., Corvelo, A., et al. (2021). Single-nucleus transcriptome analysis of human brain immune response in patients with severe COVID-19. Genome Med 13, 118. 10.1186/s13073-021-00933-8.

16. Piwecka, M., Rajewsky, N., and Rybak-Wolf, A. (2023). Single-cell and spatial transcriptomics: deciphering brain complexity in health and disease. Nat Rev Neurol 19, 346–362. 10.1038/s41582-023-00809-y.

17. Gandal, M.J., Zhang, P., Hadjimichael, E., Walker, R.L., Chen, C., Liu, S., Won, H., van Bakel, H., Varghese, M., Wang, Y., et al. (2018). Transcriptome-wide isoform-level dysregulation in ASD, schizophrenia, and bipolar disorder. Science 362, eaat8127. 10.1126/science.aat8127.

18. Hoffman, G.E., Bendl, J., Voloudakis, G., Montgomery, K.S., Sloofman, L., Wang, Y.-C., Shah, H.R., Hauberg, M.E., Johnson, J.S., Girdhar, K., et al. (2019). CommonMind Consortium provides transcriptomic and epigenomic data for Schizophrenia and Bipolar Disorder. Sci Data 6, 180. 10.1038/s41597-019-0183-6.

19. Fromer, M., Roussos, P., Sieberts, S.K., Johnson, J.S., Kavanagh, D.H., Perumal, T.M., Ruderfer, D.M., Oh, E.C., Topol, A., Shah, H.R., et al. (2016). Gene expression elucidates functional impact of polygenic risk for schizophrenia. Nat Neurosci 19, 1442–1453. 10.1038/nn.4399.

20. Kosoy, R., Fullard, J.F., Zeng, B., Bendl, J., Dong, P., Rahman, S., Kleopoulos, S.P., Shao, Z., Girdhar, K., Humphrey, J., et al. (2022). Genetics of the human microglia regulome refines Alzheimer’s disease risk loci. Nat Genet 54, 1145–1154. 10.1038/s41588-022-01149-1.

21. Young, A.M., Kumasaka, N., Calvert, F., Hammond, T.R., Knights, A., Panousis, N., Park, J.S., Schwartzentruber, J., Liu, J., Kundu, K., et al. (2021). A map of transcriptional heterogeneity and regulatory variation in human microglia. Nat Genet 53, 861–868. 10.1038/s41588-021-00875-2.

22. Gazestani, V., Kamath, T., Nadaf, N.M., Dougalis, A., Burris, S.J., Rooney, B., Junkkari, A., Vanderburg, C., Pelkonen, A., Gomez-Budia, M., et al. (2023). Early Alzheimer’s disease pathology in human cortex involves transient cell states. Cell 186, 4438–4453.e23. 10.1016/j.cell.2023.08.005.

23. Liharska, L.E., Park, Y.J., Ziafat, K., Wilkins, L., Silk, H., Linares, L.M., Thompson, R.C., Vornholt, E., Sullivan, B., Cohen, V., et al. (2023). A study of gene expression in the living human brain. Preprint at medRxiv, 10.1101/2023.04.21.23288916 10.1101/2023.04.21.23288916.

24. Pfisterer, U., Petukhov, V., Demharter, S., Meichsner, J., Thompson, J.J., Batiuk, M.Y., Asenjo-Martinez, A., Vasistha, N.A., Thakur, A., Mikkelsen, J., et al. (2020). Identification of epilepsy-associated neuronal subtypes and gene expression underlying epileptogenesis. Nat Commun 11, 5038. 10.1038/s41467-020-18752-7.

25. Storey, J.D., and Tibshirani, R. (2003). Statistical significance for genomewide studies. Proc Natl Acad Sci U S A 100, 9440–9445. 10.1073/pnas.1530509100.

26. Khatri, P., and Drăghici, S. (2005). Ontological analysis of gene expression data: current tools, limitations, and open problems. Bioinformatics 21, 3587–3595. 10.1093/bioinformatics/bti565.

27. Im, Y., and Kim, Y. (2023). A Comprehensive Overview of RNA Deconvolution Methods and Their Application. Mol Cells 46, 99–105. 10.14348/molcells.2023.2178.

28. Otero-Garcia, M., Mahajani, S.U., Wakhloo, D., Tang, W., Xue, Y.-Q., Morabito, S., Pan, J., Oberhauser, J., Madira, A.E., Shakouri, T., et al. (2022). Molecular signatures underlying neurofibrillary tangle susceptibility in Alzheimer’s disease. Neuron 110, 2929–2948.e8. 10.1016/j.neuron.2022.06.021.

29. Fromer, M., Roussos, P., Sieberts, S.K., Johnson, J.S., Kavanagh, D.H., Perumal, T.M., Ruderfer, D.M., Oh, E.C., Topol, A., Shah, H.R., et al. (2016). Gene expression elucidates functional impact of polygenic risk for schizophrenia. Nat Neurosci 19, 1442–1453. 10.1038/nn.4399.

30. Hoffman, G.E., Bendl, J., Voloudakis, G., Montgomery, K.S., Sloofman, L., Wang, Y.-C., Shah, H.R., Hauberg, M.E., Johnson, J.S., Girdhar, K., et al. (2019). CommonMind Consortium provides transcriptomic and epigenomic data for Schizophrenia and Bipolar Disorder. Sci Data 6, 180. 10.1038/s41597-019-0183-6.

31. Bakken, T.E., Jorstad, N.L., Hu, Q., Lake, B.B., Tian, W., Kalmbach, B.E., Crow, M., Hodge, R.D., Krienen, F.M., Sorensen, S.A., et al. (2021). Comparative cellular analysis of motor cortex in human, marmoset and mouse. Nature 598, 111–119. 10.1038/s41586-021-03465-8.

32. Kamath, T., Abdulraouf, A., Burris, S.J., Langlieb, J., Gazestani, V., Nadaf, N.M., Balderrama, K., Vanderburg, C., and Macosko, E.Z. (2022). Single-cell genomic profiling of human dopamine neurons identifies a population that selectively degenerates in Parkinson’s disease. Nat Neurosci 25, 588–595. 10.1038/s41593-022-01061-1.

33. Velmeshev, D., Schirmer, L., Jung, D., Haeussler, M., Perez, Y., Mayer, S., Bhaduri, A., Goyal, N., Rowitch, D.H., and Kriegstein, A.R. (2019). Single-cell genomics identifies cell type-specific molecular changes in autism. Science 364, 685–689. 10.1126/science.aav8130.

34. Jäkel, S., Agirre, E., Mendanha Falcão, A., van Bruggen, D., Lee, K.W., Knuesel, I., Malhotra, D., Ffrench-Constant, C., Williams, A., and Castelo-Branco, G. (2019). Altered human oligodendrocyte heterogeneity in multiple sclerosis. Nature 566, 543–547. 10.1038/s41586-019-0903-2.

35. Lau, S.-F., Cao, H., Fu, A.K.Y., and Ip, N.Y. (2020). Single-nucleus transcriptome analysis reveals dysregulation of angiogenic endothelial cells and neuroprotective glia in Alzheimer’s disease. Proc Natl Acad Sci U S A 117, 25800–25809. 10.1073/pnas.2008762117.

36. Gazestani, V., Kamath, T., Nadaf, N.M., Dougalis, A., Burris, S.J., Rooney, B., Junkkari, A., Vanderburg, C., Pelkonen, A., Gomez-Budia, M., et al. (2023). Early Alzheimer’s disease pathology in human cortex involves transient cell states. Cell 186, 4438–4453.e23. 10.1016/j.cell.2023.08.005.

37. Williams, S.E., Mealer, R.G., Scolnick, E.M., Smoller, J.W., and Cummings, R.D. (2020). Aberrant glycosylation in schizophrenia: a review of 25 years of post-mortem brain studies. Mol Psychiatry 25, 3198–3207. 10.1038/s41380-020-0761-1.

38. Vornholt, E., Luo, D., Qiu, W., McMichael, G.O., Liu, Y., Gillespie, N., Ma, C., and Vladimirov, V.I. (2019). Postmortem brain tissue as an underutilized resource to study the molecular pathology of neuropsychiatric disorders across different ethnic populations. Neuroscience & Biobehavioral Reviews 102, 195–207. 10.1016/j.neubiorev.2019.04.015.

39. Liu, S.-H., Du, Y., Chen, L., and Cheng, Y. (2022). Glial Cell Abnormalities in Major Psychiatric Diseases: A Systematic Review of Postmortem Brain Studies. Mol Neurobiol 59, 1665–1692. 10.1007/s12035-021-02672-8.

40. Leung, E., Lau, E.W., Liang, A., de Dios, C., Suchting, R., Östlundh, L., Masdeu, J.C., Fujita, M., Sanches, M., Soares, J.C., et al. (2022). Alterations in brain synaptic proteins and mRNAs in mood disorders: a systematic review and meta-analysis of postmortem brain studies. Mol Psychiatry 27, 1362–1372. 10.1038/s41380-021-01410-9.

41. Sandroni, C., Cronberg, T., and Sekhon, M. (2021). Brain injury after cardiac arrest: pathophysiology, treatment, and prognosis. Intensive Care Med 47, 1393–1414. 10.1007/s00134-021-06548-2.

42. Amantea, D., Micieli, G., Tassorelli, C., Cuartero, M.I., Ballesteros, I., Certo, M., Moro, M.A., Lizasoain, I., and Bagetta, G. (2015). Rational modulation of the innate immune system for neuroprotection in ischemic stroke. Frontiers in Neuroscience 9. 10.3389/fnins.2015.00147.

43. Jayaraj, R.L., Azimullah, S., Beiram, R., Jalal, F.Y., and Rosenberg, G.A. (2019). Neuroinflammation: friend and foe for ischemic stroke. Journal of Neuroinflammation 16, 142. 10.1186/s12974-019-1516-2.

44. Marques, B.L., Carvalho, G.A., Freitas, E.M.M., Chiareli, R.A., Barbosa, T.G., Di Araújo, A.G.P., Nogueira, Y.L., Ribeiro, R.I., Parreira, R.C., Vieira, M.S., et al. (2019). The role of neurogenesis in neurorepair after ischemic stroke. Seminars in Cell & Developmental Biology 95, 98–110. 10.1016/j.semcdb.2018.12.003.

45. Kreuzberg, M., Kanov, E., Timofeev, O., Schwaninger, M., Monyer, H., and Khodosevich, K. (2010). Increased subventricular zone-derived cortical neurogenesis after ischemic lesion. Experimental Neurology 226, 90–99. 10.1016/j.expneurol.2010.08.006.

46. Regulation of transcription factors by neuronal activity | Nature Reviews Neuroscience https://www.nature.com/articles/nrn987.

47. Takahashi, H., Yamamoto, T., and Tsuboi, A. (2023). Molecular mechanisms underlying activity-dependent ischemic tolerance in the brain. Neuroscience Research 186, 3–9. 10.1016/j.neures.2022.10.005.

48. White, B.C., Sullivan, J.M., DeGracia, D.J., O’Neil, B.J., Neumar, R.W., Grossman, L.I., Rafols, J.A., and Krause, G.S. (2000). Brain ischemia and reperfusion: molecular mechanisms of neuronal injury. Journal of the Neurological Sciences 179, 1–33. 10.1016/S0022-510X(00)00386-5.

49. Gurina, T.S., and Mohiuddin, S.S. (2023). Biochemistry, Protein Catabolism. In StatPearls (StatPearls Publishing).

50. Mukandala, G., Tynan, R., Lanigan, S., and O’Connor, J.J. (2016). The Effects of Hypoxia and Inflammation on Synaptic Signaling in the CNS. Brain Sci 6, 6. 10.3390/brainsci6010006.

51. Dent, K.A., Christie, K.J., Bye, N., Basrai, H.S., Turbic, A., Habgood, M., Cate, H.S., and Turnley, A.M. (2015). Oligodendrocyte Birth and Death following Traumatic Brain Injury in Adult Mice. PLoS One 10, e0121541. 10.1371/journal.pone.0121541.

52. McTigue, D.M., and Tripathi, R.B. (2008). The life, death, and replacement of oligodendrocytes in the adult CNS. Journal of Neurochemistry 107, 1–19. 10.1111/j.1471-4159.2008.05570.x.

53. Chen, Y., Yi, Q., Liu, G., Shen, X., Xuan, L., and Tian, Y. (2013). Cerebral white matter injury and damage to myelin sheath following whole-brain ischemia. Brain Research 1495, 11–17. 10.1016/j.brainres.2012.12.006.

54. Stadelmann, C., Timmler, S., Barrantes-Freer, A., and Simons, M. (2019). Myelin in the Central Nervous System: Structure, Function, and Pathology. Physiological Reviews 99, 1381–1431. 10.1152/physrev.00031.2018.

55. Miller, M.J., Kangas, C.D., and Macklin, W.B. (2009). Neuronal expression of the proteolipid protein gene in the medulla of the mouse. J Neurosci Res 87, 2842–2853. 10.1002/jnr.22121.

56. Sarret, C., Combes, P., Micheau, P., Gelot, A., Boespflug-Tanguy, O., and Vaurs-Barriere, C. (2010). Novel neuronal proteolipid protein isoforms encoded by the human myelin proteolipid protein 1 gene. Neuroscience 166, 522–538. 10.1016/j.neuroscience.2009.12.047.

57. Will, C.L., and Lührmann, R. (2011). Spliceosome Structure and Function. Cold Spring Harb Perspect Biol 3, a003707. 10.1101/cshperspect.a003707.

58. Guo, H., Xia, L., Wang, W., Xu, W., Shen, X., Wu, X., He, T., Jiang, X., Xu, Y., Zhao, P., et al. (2023). Hypoxia induces alterations in tRNA modifications involved in translational control. BMC Biol 21, 39. 10.1186/s12915-023-01537-x.

59. Blaze, J., and Akbarian, S. (2022). The tRNA regulome in neurodevelopmental and neuropsychiatric disease. Mol Psychiatry 27, 3204–3213. 10.1038/s41380-022-01585-9.

60. Zeisel, A., Muñoz-Manchado, A.B., Codeluppi, S., Lönnerberg, P., La Manno, G., Juréus, A., Marques, S., Munguba, H., He, L., Betsholtz, C., et al. (2015). Cell types in the mouse cortex and hippocampus revealed by single-cell RNA-seq. Science 347, 1138–1142. 10.1126/science.aaa1934.

61. Darmanis, S., Sloan, S.A., Zhang, Y., Enge, M., Caneda, C., Shuer, L.M., Hayden Gephart, M.G., Barres, B.A., and Quake, S.R. (2015). A survey of human brain transcriptome diversity at the single cell level. Proceedings of the National Academy of Sciences 112, 7285–7290. 10.1073/pnas.1507125112.

62. Chen, B., Khodadoust, M.S., Liu, C.L., Newman, A.M., and Alizadeh, A.A. (2018). Profiling tumor infiltrating immune cells with CIBERSORT. Methods Mol Biol 1711, 243–259. 10.1007/978-1-4939-7493-1_12.

63. Steen, C.B., Liu, C.L., Alizadeh, A.A., and Newman, A.M. (2020). Profiling Cell Type Abundance and Expression in Bulk Tissues with CIBERSORTx. Methods Mol Biol 2117, 135–157. 10.1007/978-1-0716-0301-7_7.

64. Menden, K., Marouf, M., Oller, S., Dalmia, A., Magruder, D.S., Kloiber, K., Heutink, P., and Bonn, S. (2020). Deep learning–based cell composition analysis from tissue expression profiles. Science Advances 6, eaba2619. 10.1126/sciadv.aba2619.

65. Pavlou, M., Ambler, G., Seaman, S.R., Guttmann, O., Elliott, P., King, M., and Omar, R.Z. (2015). How to develop a more accurate risk prediction model when there are few events. BMJ 351, h3868. 10.1136/bmj.h3868.

66. Kruppa, J., Ziegler, A., and König, I.R. (2012). Risk estimation and risk prediction using machine-learning methods. Hum Genet 131, 1639–1654. 10.1007/s00439-012-1194-y.

67. Siletti, K., Hodge, R., Mossi Albiach, A., Lee, K.W., Ding, S.-L., Hu, L., Lönnerberg, P., Bakken, T., Casper, T., Clark, M., et al. (2023). Transcriptomic diversity of cell types across the adult human brain. Science 382, eadd7046. 10.1126/science.add7046.

68. Ramirez, E.P.C., Keller, C.E., and Vonsattel, J.P. (2018). The New York Brain Bank of Columbia University: practical highlights of 35 years of experience. Handb Clin Neurol 150, 105–118. 10.1016/B978-0-444-63639-3.00008-6.

69. Vonsattel, J.P.G., Amaya, M.D.P., Cortes, E.P., Mancevska, K., and Keller, C.E. (2008). Twenty-first century brain banking: practical prerequisites and lessons from the past: the experience of New York Brain Bank, Taub Institute, Columbia University. Cell Tissue Banking 9, 247–258. 10.1007/s10561-008-9079-y.

70. Lun, A.T.L., Riesenfeld, S., Andrews, T., Dao, T.P., Gomes, T., Marioni, J.C., and participants in the 1st Human Cell Atlas Jamboree (2019). EmptyDrops: distinguishing cells from empty droplets in droplet-based single-cell RNA sequencing data. Genome Biology 20, 63. 10.1186/s13059-019-1662-y.

71. Young, M.D., and Behjati, S. (2020). SoupX removes ambient RNA contamination from droplet-based single-cell RNA sequencing data. GigaScience 9, giaa151. 10.1093/gigascience/giaa151.

72. Stuart, T., Butler, A., Hoffman, P., Hafemeister, C., Papalexi, E., Mauck, W.M., Hao, Y., Stoeckius, M., Smibert, P., and Satija, R. (2019). Comprehensive Integration of Single-Cell Data. Cell 177, 1888–1902.e21. 10.1016/j.cell.2019.05.031.

73. Luecken, M.D., and Theis, F.J. (2019). Current best practices in single-cell RNA-seq analysis: a tutorial. Molecular Systems Biology 15, e8746. 10.15252/msb.20188746.

74. Germain, P.-L., Lun, A., Garcia Meixide, C., Macnair, W., and Robinson, M.D. (2022). Doublet identification in single-cell sequencing data using scDblFinder. F1000Res 10, 979. 10.12688/f1000research.73600.2.

75. Korsunsky, I., Millard, N., Fan, J., Slowikowski, K., Zhang, F., Wei, K., Baglaenko, Y., Brenner, M., Loh, P., and Raychaudhuri, S. (2019). Fast, sensitive and accurate integration of single-cell data with Harmony. Nat Methods 16, 1289–1296. 10.1038/s41592-019-0619-0.

76. Tirosh, I., Izar, B., Prakadan, S.M., Wadsworth, M.H., Treacy, D., Trombetta, J.J., Rotem, A., Rodman, C., Lian, C., Murphy, G., et al. (2016). Dissecting the multicellular ecosystem of metastatic melanoma by single-cell RNA-seq. Science 352, 189–196. 10.1126/science.aad0501.

77. McCarthy, D.J., Campbell, K.R., Lun, A.T.L., and Wills, Q.F. (2017). Scater: pre-processing, quality control, normalization and visualization of single-cell RNA-seq data in R. Bioinformatics 33, 1179–1186. 10.1093/bioinformatics/btw777.

78. Robinson, M.D., McCarthy, D.J., and Smyth, G.K. (2010). edgeR: a Bioconductor package for differential expression analysis of digital gene expression data. Bioinformatics 26, 139–140. 10.1093/bioinformatics/btp616.

79. Hoffman, G.E., and Schadt, E.E. (2016). variancePartition: interpreting drivers of variation in complex gene expression studies. BMC Bioinformatics 17, 483. 10.1186/s12859-016-1323-z.

80. Law, C.W., Chen, Y., Shi, W., and Smyth, G.K. (2014). voom: precision weights unlock linear model analysis tools for RNA-seq read counts. Genome Biology 15, R29. 10.1186/gb-2014-15-2-r29.

81. Benjamini, Y., and Hochberg, Y. (1995). Controlling the False Discovery Rate: A Practical and Powerful Approach to Multiple Testing. Journal of the Royal Statistical Society. Series B (Methodological) 57, 289–300.

82. The Gene Ontology Consortium, Aleksander, S.A., Balhoff, J., Carbon, S., Cherry, J.M., Drabkin, H.J., Ebert, D., Feuermann, M., Gaudet, P., Harris, N.L., et al. (2023). The Gene Ontology knowledgebase in 2023. Genetics 224, iyad031. 10.1093/genetics/iyad031.

83. Alexa, A., and Rahnenfuhrer, J. Gene set enrichment analysis with topGO.

84. Ritchie, M.E., Phipson, B., Wu, D., Hu, Y., Law, C.W., Shi, W., and Smyth, G.K. (2015). limma powers differential expression analyses for RNA-sequencing and microarray studies. Nucleic Acids Research 43, e47. 10.1093/nar/gkv007.

85. Zou, H., and Hastie, T. (2005). Regularization and Variable Selection Via the Elastic Net. Journal of the Royal Statistical Society Series B: Statistical Methodology 67, 301–320. 10.1111/j.1467-9868.2005.00503.x.

86. Sing, T., Sander, O., Beerenwinkel, N., and Lengauer, T. (2005). ROCR: visualizing classifier performance in R. Bioinformatics 21, 3940–3941. 10.1093/bioinformatics/bti623.

