## Supplemental Figures for "Characterizing cell type specific transcriptional differences between the living and postmortem human brain"

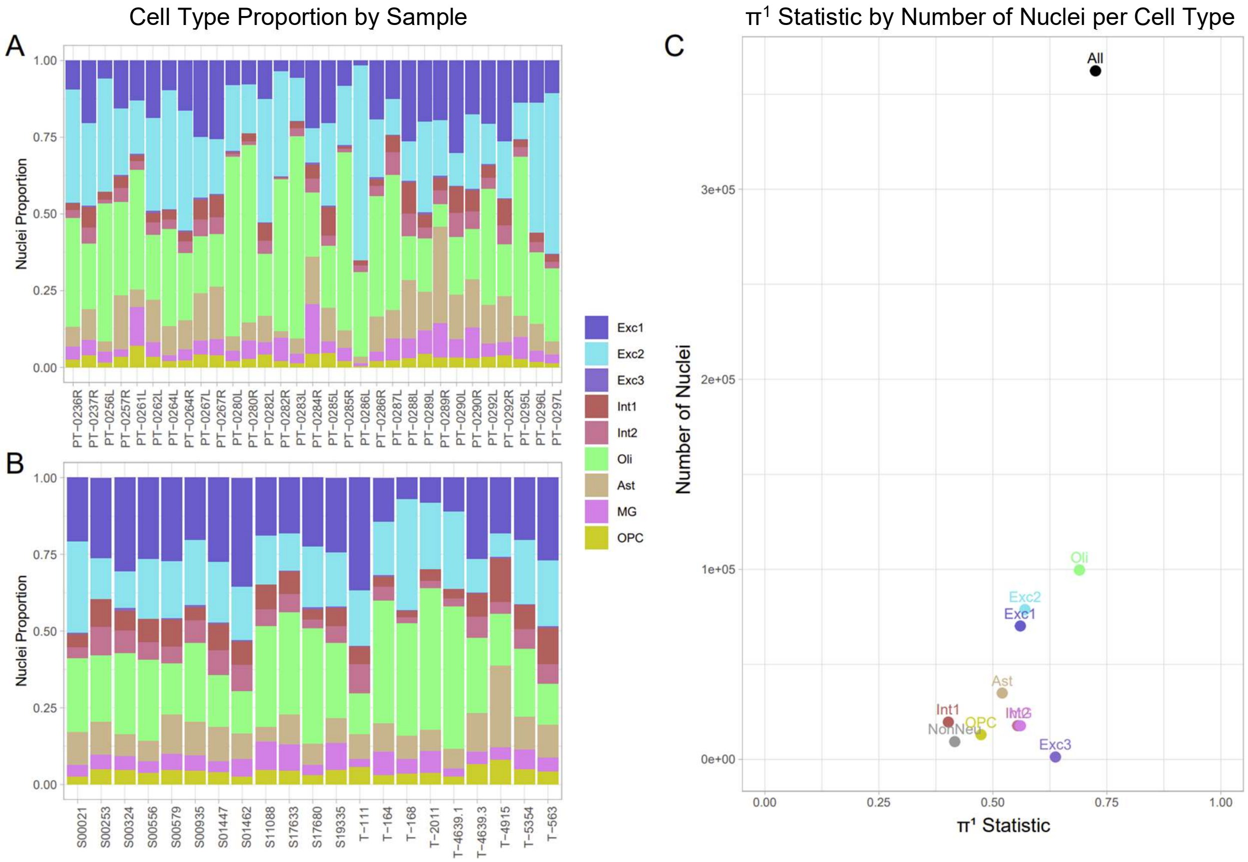


***Supplemental Figure 1*: Cell type proportions by sample and π[1] estimates across differential expression (DE) analyses. – A)** The proportion (y-axis) of each annotated cell type (colors) across samples (x-axis) obtained from living individuals (LIV). **B)** The proportion (y-axis) of each annotated cell type (colors) across postmortem samples (PM, x-axis). **C)** Power estimates of significant associations ($\pi^1$; x-axis) as a function of the number of nuclei per cell type (y-axis). Colors is defined in the legend of panels A and B and cell types are defined above each point.

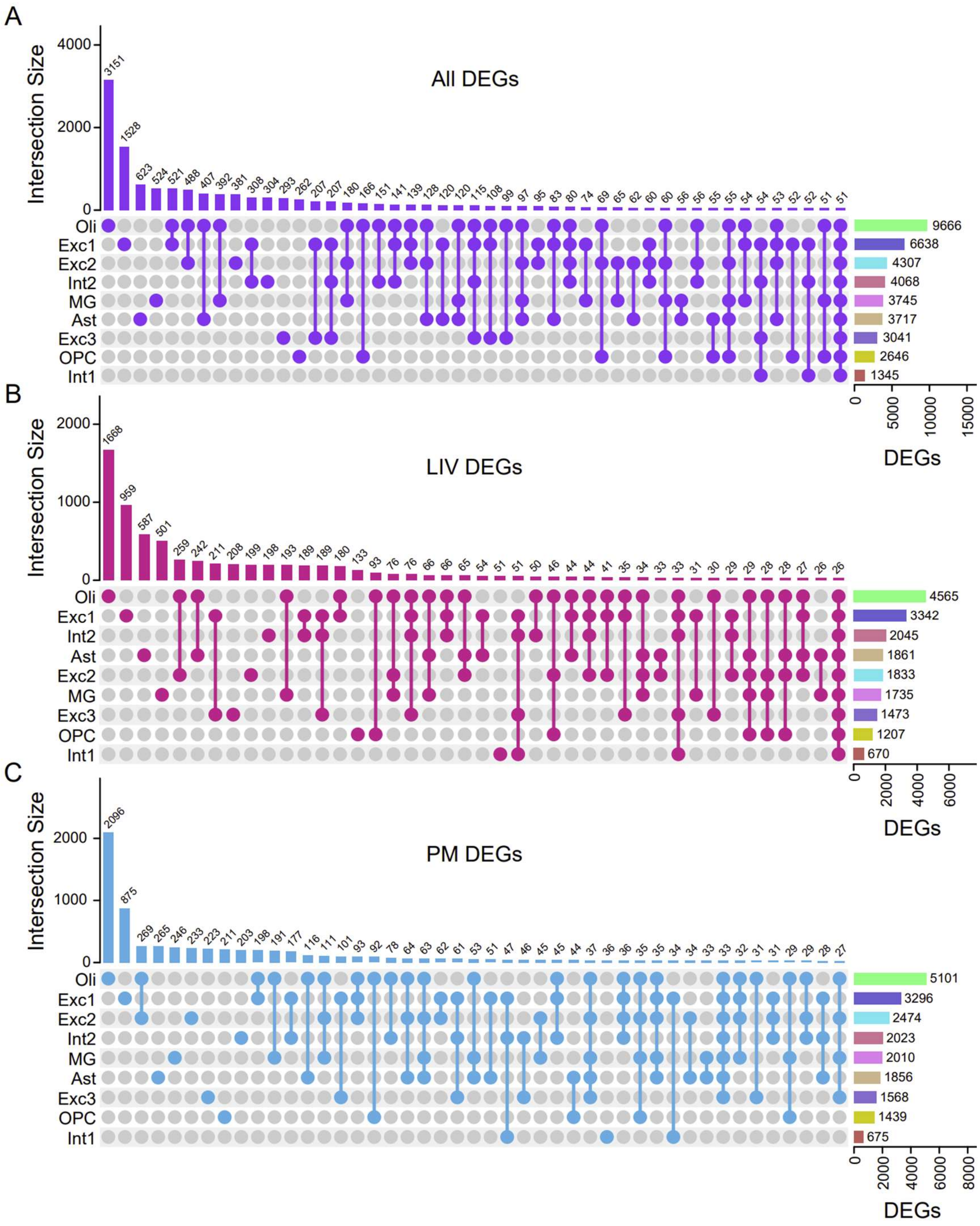


***Supplemental Figure 2:*** **Overlap of DEGs across cell types –** Upset plots presenting the groups of overlapping DEG sets (intersects) presented for all DEGs (**A**, sets with > 50 genes only), LIV DEGs (**B**, sets with > 25 genes only), and PM DEGs (**C**, sets with > 25 genes only). Intersects are presented by the colored points and lines connecting all cell types in the overlapping set in the bar above. The y-axis and vertical bars are the number of genes in each intersects defined by the dotted line below. Horizontal bars represent the number of total DEGs for a given cell type (number specified on each bar; x-axis). Cell types present in each intersect are defined on the right of the points.

A

| | LBP *n=52* (discovery) | Hodge et al. *n=8* (replication) |
|---|---|---|
| **# Cells** | 108,875<br>*LIV*=33,644<br>*PM*=75,231 | 15,005<br>*LIV*=711<br>*PM*=14,294 |
| **Sequencing** | 10X | SMART-seq |
| **Brain region** | Prefrontal Cortex (PFC) | Middle temporal gyrus (MTG) |
| **Phenotype** | PD/MDD/OCD | Epilepsy |
| **Cell sorting** | None | NeuN FACS sorting |

B

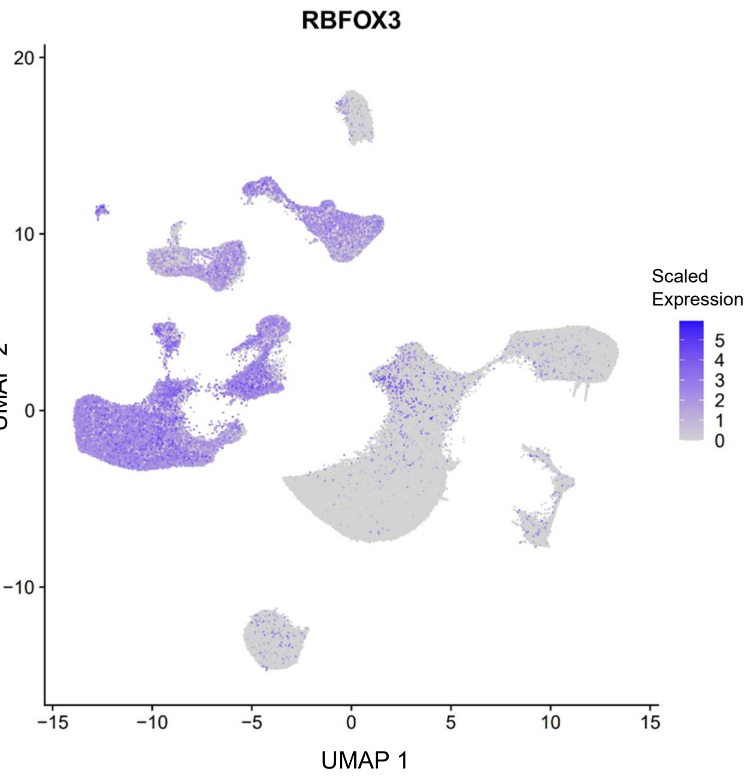


***Supplemental Figure 3:*** **Hodge et al. external replication supplement** – **A)** Table of key differences in study design between the Living Brain Project (LBP) discovery and Hodge et al. replication dataset. **B)** UMAP of RBFOX3 activity, presented as a gradient from grey (low expression) to blue (high gene expression) across all nuclei highlighting the neuronal clusters representative of NeuN+ neuron in the LBP dataset based on their. The x and y-axes are the UMAP coordinates.

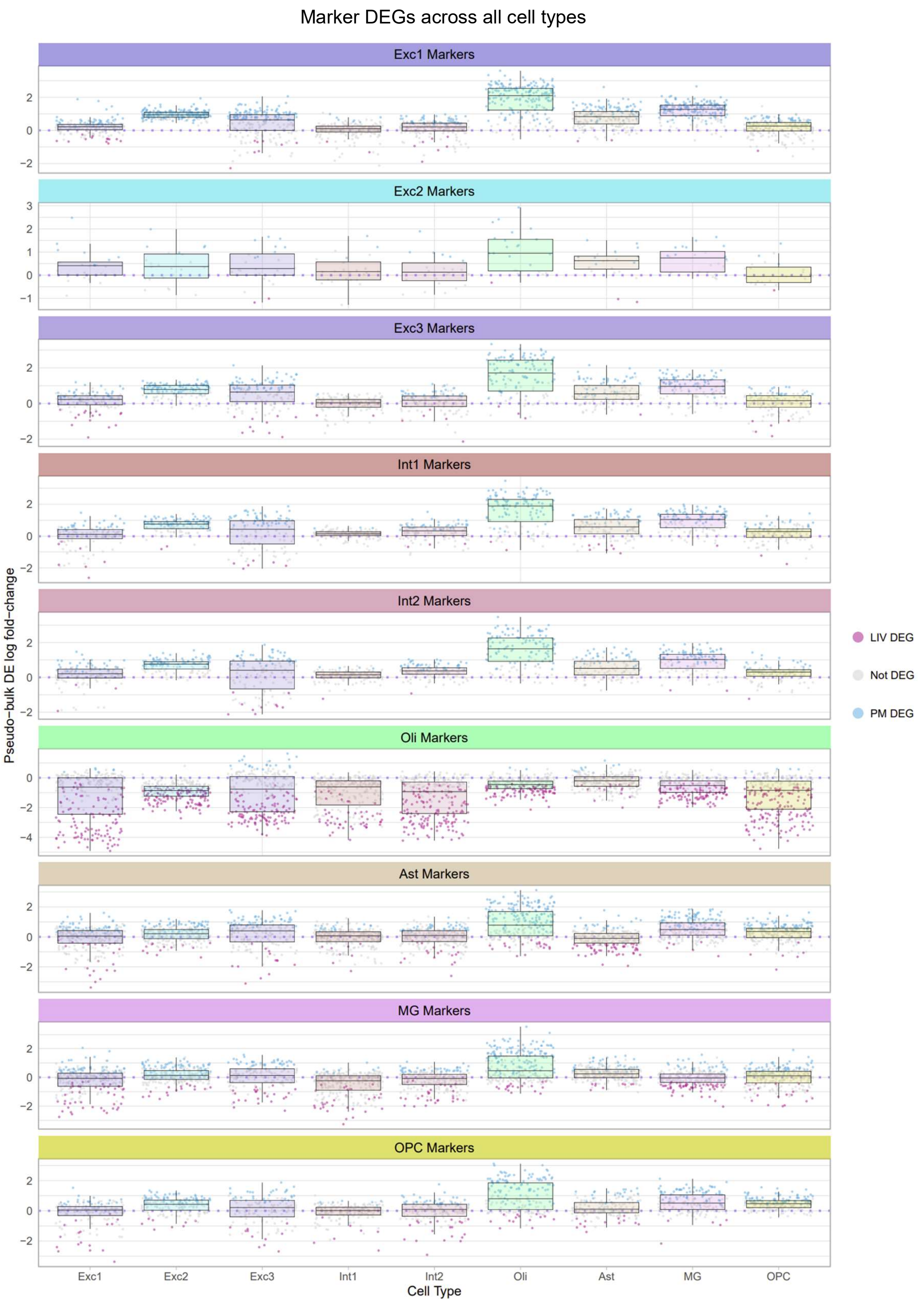


***Supplemental Figure 4:*** **Marker DEGs across all cell types –** DEG classification (LIV DEG = increased transcript abundance in LIV samples (pink points), PM DEG = increased transcript. The x and y axes are the logFC of the DE analyses and the cell type in which DE analysis is performed respectively. Each facet represents a different set of cell type markers.

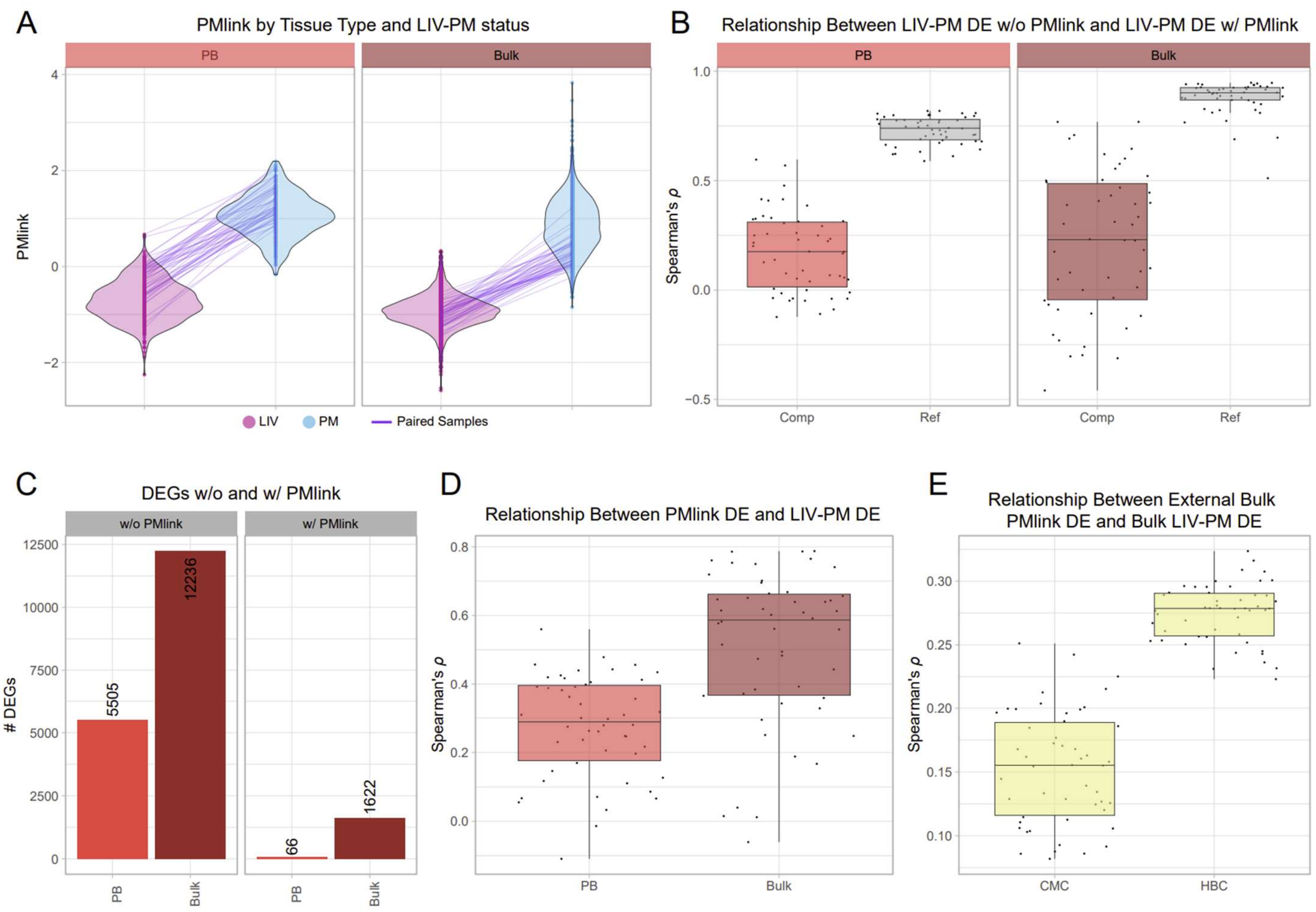


***Supplemental Figure 5:*** **All-nuclei pseudo-bulk and bulk PMlink results** – **A)** Violin plots presenting the distribution of PMlink scores (y-axis) calculated from the scaled postmortem probability across all 50 elastic-net iterations separated by the LIV-PM status of samples (LIV=pink, PM=blue). Each point represents a sample also colored by LIV-PM status with paired samples within the same testing set connected by a purple line. The violin plots are faceted for PMlink scores calculated on all-nuclei pseudo-bulk and bulk. **B)** Spearman's correlation coefficients (ρ; y-axis) comparing the logFCs from LIV-PM DE calculated on the testing set before and after including PMlink in the linear model. Points represent the ρ for each testing sample split iteration faceted by tissue type in which PMlink is calculated. Within each facet, the colored boxplot represents the with and without PMlink comparison (Comp; x-axis) and the grey boxplot represents the reference comparison the testing set LIV-PM DE without PMlink and the holdout set LIV-PM DE (Ref; x-axis). **C)** Bar plots depicting the average number of significant DEGs (FDR $\leq$ 0.05) across testing set LIV-PM DE analyses before (left) and after (right) including PMlink in the simple linear model across both tissue types (x-axis). **D)** Spearman's correlation coefficients (ρ; y-axis) comparing the logFCs from LIV-PM DE calculated on the holdout set and PMlink DE calculated on the postmortem subset of each testing set. Points represent the ρ for each testing sample split iteration with the x-axis and colors representing the tissue type each comparison was performed in. **E)** Spearman's correlation coefficients (ρ; y-axis) comparing the logFCs from the external bulk (CMC and HBC) PMlink DE and holdout LIV-PM DE with each point representing a single ρ.
